# β-Amyloid as a new target to suppress tonic PTH hypersecretion in primary hyperparathyroidism

**DOI:** 10.1101/2025.05.27.25328314

**Authors:** Chia-Ling Tu, Zhiqiang Cheng, Nicholas Szeto, Sofya Savransky, Timofey Glinin, Dolores M. Shoback, Julie A. Sosa, Jean-Pierre Vilardaga, James Koh, Wenhan Chang

## Abstract

Primary hyperparathyroidism (PHPT) is a common endocrine disorder of aging closely linked to vitamin D deficiency. Reduced vitamin D receptor activities promote parathyroid hormone (PTH) hypersecretion by increasing the heterodimerization of the type B γ-aminobutyric acid receptor 1 (GABA_B1_R) with the extracellular Ca^2+^-sensing receptor (CaSR) in parathyroid cells; however, endogenous activators of the heterodimers are unknown. Here we uncovered increased expression of the β-amyloid peptide (Aβ_42_) cleaved from the amyloid precursor protein in parathyroid cells from PHPT patients and aging mice, and the ability of exogenous Aβ_42_ to promote tonic PTH secretion from murine or human parathyroid glands *ex vivo*. Conversely, parathyroid-specific *App* gene knockout reduced tonic PTH secretion and lowered serum PTH levels in mice. The absence of an Aβ_42_ effect on PTH secretion in parathyroid glands lacking CaSR or GABA_B1_R supports direct interactions between Aβ_42_ and the heterodimer. *In situ* proteomic profiling of parathyroid glands from PHPT patients closely correlated lower serum 25-hydroxyvitamin D levels with increased GABA_B1_R /CaSR heterodimer expression, β-amyloidogenesis, and phosphorylation of Tau, a downstream effector of Aβ_42_. Concurrent ablation of *App* or the Tau-encoding *Mapt* gene prevented tonic PTH hypersecretion in parathyroid-specific *Vdr*-KO mice. Likewise, weekly administration of an Aβ_42_-neutralizing antibody suppressed tonic PTH hypersecretion and synergized with daily administration of cinacalcet, a calcimimetic that activates CaSR homodimers, to reduce serum PTH levels in aging-induced hyperparathyroidism (HPT) mice. These data demonstrated novel functions of Aβ_42_ in driving tonic PTH secretion by activating GABA_B1_R/CaSR heterodimers and suggest the potential for targeting Aβ_42_ in PHPT treatment.

**One Sentence Summary:** Blocking aberrant signaling through Aβ, the GABA_B1_R/CaSR dimer, and Tau can suppress tonic PTH hypersecretion in hyperparathyroidism associated with vitamin D deficiency.

## INTRODUCTION

PHPT disproportionately affects the elderly and can cause a spectrum of debilitating sequelae in bone, kidney, the cardiovascular system, brain, and many other organs (*1, 2*). In the U.S., the PHPT incidence of 15-30 persons per 100,000 continues trending upwards as the nation’s median population ages (*3*). Severe osteoporosis, nephrolithiasis, hypertension, and neurological deficits secondary to PHPT can be challenging to manage (*4–6*). Dietary supplementation with calcium and vitamin D has been shown to reduce PHPT symptoms although efficacy varies substantially, possibly due to multiple confounders including genetics, age, and sex, and phenotypic subtypes of PHPT tumors with distinct secretory properties (*7*). Thus, an in-depth mechanistic understanding of PHPT pathology, particularly pertaining to actions of vitamin D, is essential to improve the specificity and precision of pre-operative counseling and post-surgical management and to develop alternative treatment options for patients who are non-surgical candidates.

Parathyroid glands critically maintain systemic Ca^2+^ homeostasis by regulating PTH secretion in response to acute changes in serum Ca^2+^ levels signaled *via* the CaSR and sustained changes in vitamin D status *via* the vitamin D receptor (VDR). 1,25-dihydroxyvitamin D [1,25(OH)_2_D], the bioactive metabolite converted from circulating 25-hydroxyvitamin D (25OHD), is thought to prevent chronic PTH hypersecretion and the eventual development of hyperparathyroidism by stimulating *CASR* gene transcription, inhibiting *PTH* gene transcription, and blocking parathyroid cell proliferation (*8*). PHPT severity often correlates inversely with serum 25OHD levels in human patients (*9*), but the molecular mechanisms coupling VDR signaling to suppression of PTH secretion remain undefined. The current paradigm proposes that the impaired ability of PHPT tumors to suppress PTH secretion in response to rising extracellular [Ca^2+^] ([Ca^2+^]_e_) is simply a consequence of reduced levels of homomeric CaSR complexes due to suppressed *CASR* gene transcription caused by vitamin D deficiency or aging (*8*). However, this paradigm fails to explain intrinsic tonic PTH secretion at low [Ca^2+^]_e_ under normal physiological conditions, when homomeric CaSRs are inactive or why, in many cases of PHPT, the causative parathyroid tumors retain normal Ca^2+^-responsiveness despite their elevated tonic PTH production (*7*). To address these questions, we sought to identify mechanisms that can constitutively raise tonic PTH secretion in PHPT.

We showed previously that reduced CaSR expression and increased GABA_B1_R (encoded by *Gabbr1* gene) expression in the parathyroid glands of patients with primary or secondary hyperparathyroidism augmented heterodimerization of the two receptors and increased tonic PTH secretion (*10*). Parathyroid-specific *Gabbr1* gene KO to block formation of GABA_B1_R/CaSR heterodimers lowered serum PTH levels by reducing the maximal PTH secretion rate (PTH_Max_) and the Ca^2+^-setpoint (defined as [Ca^2+^]_e_ required to suppress 50% of Ca^2+^-suppressible PTH secretion) of the parathyroid glands in normal mice or mouse models of hereditary PHPT or chronic Ca^2+^ insufficiency (*10*). Conversely, GABA_B1_R agonists (e.g., baclofen and GABA) enhanced PTH_Max_ (tonic PTH secretion) and/or raised the Ca^2+^-setpoint (i.e., decreased Ca^2+^-sensitivity) (*10*). Since the agonistic effects of baclofen depended on the co-expression of GABA_B1_R and CaSR (*10*), the impact of GABA_B1_R on tonic PTH secretion and Ca^2+^-responsiveness likely required heterodimerization of these two receptors to antagonize G_i_ and G_q_ signaling of the homodimeric CaSR and perhaps other unknown G protein-coupled signaling molecules. Parathyroid-specific KO of glutamate decarboxylase 1 and 2, two GABA-generating enzymes, decreased the Ca^2+^-setpoint (enhancing Ca^2+^-responsiveness) without significantly altering tonic PTH secretion (*10*). While these results demonstrated paracrine/autocrine actions of endogenic GABA in regulating parathyroid cell Ca^2+^-sensitivity, the inability of endogenous GABA blockade to affect PTH_Max_ indicates the presence of additional activators of the GABA_B1_R/CaSR heterodimer that mediate tonic PTH secretion.

*Rice et al.* reported that the cleavage products of the β-amyloid precursor protein (APP) may serve as an allosteric agonist of the GABA_B1_R/GABA_B2_R heterodimer (*11*). As initial steps of APP proteolysis, the α- or β-secretase catalyzes the release of secreted forms of APP ectodomain (sAPP-α or sAPP-β, respectively), which contain a 17-amino acid extension domain (ExtD) that binds and activates the sushi 1 domain of the GABA_B1_R (*11*). In the “pathogenic” β-amyloidogenic mode, sequential cleavages at sites in the APP ectodomain by β-secretase and in the membrane-spanning domain by γ-secretase produce the Aβ moiety, which are predominantly 36 to 42 amino acid peptides (Aβ_36_ to Aβ_42_) in monomeric, oligomeric, or fibrillar forms. Excessive Aβ_42_ buildup is linked to hyperphosphorylation of the microtubule-associated protein, Tau, a critical initiator in the neurodegenerative process of Alzheimer’s disease (AD) (*12, 13*). While fibrillar Aβ_42_ is prone to form Aβ_42_-enriched plaques frequently seen in the brains of AD patients, soluble Aβ_42_ oligomers likely act directly on membrane receptors in neural cells (*14*); however, the identities of Aβ_42_ receptor(s) remain to be defined. Soluble Aβ_42_ has been shown to stimulate Ca^2+^-permeable nonselective cation channels in hippocampal neurons (*15*), induce intracellular accumulation of endogenous Aβ in neurons and astrocytes (*16*), and promote Tau hyperphosphorylation (*16–18*) in a CaSR-dependent manner. Based on these findings, we hypothesized that proteolytic cleavage products of APP could act as alternative ligands of the GABA_B1_R/CaSR heterodimer in sustaining tonic PTH secretion. We evaluated the expression of APP, Aβ_42_, and APP proteolytic processing enzymes in parathyroid adenomas from PHPT patients, particularly those who presented with pre-operative vitamin D-deficiency. We employed parathyroid-specific gene ablation and *ex vivo* interrogative assays to examine the actions of APP/Aβ_42_ in driving tonic PTH secretion, defined the interactions of the Aβ_42_ peptide with CaSR and GABA_B1_R, and studied the impact of VDR signaling on the APP/Aβ_42_/Tau signaling cascade in the context of PHPT and aging. Finally, we compared the actions and efficacy of an Aβ_42_-neutralizing antibody, a murine version of aducanumab (mAdu) (*19*), *versus* cinacalcet, a clinically employed calcimimetic, on suppressing PTH hypersecretion and serum PTH levels in aging mice with HPT.

## RESULTS

### Distinct patterns of PTH hypersecretion in aging, vitamin D deficiency, and CaSR insufficiency

Aging, vitamin D deficiency, and hereditary reduction of CaSR expression/activity are causally linked to hyperparathyroidism, but how these conditions alter PTH secretory dynamics has not been fully characterized. We compared PTH secretory responses to increasing [Ca^2+^]_e_ in *ex vivo* PTGs cultured from both mature and aging C57/BL6 mice, as well as from mature ^PTC^*Vdr*^βflox/βflox^ (^PTC^*Vdr*^-/-^) and ^PTC^*Casr*^wt/Δflox^ (^PTC^*Casr*^+/-^) mice. The latter 2 groups manifested parathyroid-specific homozygous VDR KO and heterozygous CaSR KO to mimic vitamin D deficiency and CaSR insufficiency, respectively.

Parathyroid glands from mice at 24 versus 6 months of age (MOA) showed a significant increase in tonic PTH secretion, with a >100% increase in PTH_Max_ and reduced Ca^2+^-sensitivity indicated by a right-shifted Ca^2+^-setpoint by ≈0.35 mM (**Fig. 1A**). These altered secretory properties resulted in age-dependent elevations of serum PTH (sPTH) (**Fig. 1B**) and total calcium (sCa) (**Fig. 1C**) levels from 3 to 6 and 24 MOA. Parathyroid glands cultured from the ^PTC^*Vdr*^-/-^ mice at 3 MOA displayed an ≈100% increase in PTH_Max_ without significantly altering the Ca^2+^- setpoint (**Fig. 1D**), while parathyroid glands cultured from the ^PTC^*Casr*^+/-^ mice manifested a profound right-shift (≈0.4 mM) in the Ca^2+^-setpoint with a smaller (≈40%) increase in PTH_Max_ (**Fig. 1D**). Remarkably, the latter PTH_max_ effects in ^PTC^*Casr*^+/-^ parathyroid glands were further augmented by the concurrent ablation of the *Vdr* gene in the ^PTC^*Casr*^+/-^*Vdr*^-/-^ mice (**Fig. 1D**), revealing a CaSR-independent action of VDR in suppressing tonic PTH secretion. This role for VDR was further corroborated *in vivo* by the ability of concurrent *Vdr* KO to triple or double the effect of *Casr* gene knockdown on sPTH levels in female (**Fig. 1E**) and male (**fig. S1A**) mice, without further raising sCa levels (**Fig. 1F** and **fig. S1B**). These data demonstrate two distinct regulatory modes of PTH secretion: (i) tonic secretion dependent on VDR signaling, and (ii) [Ca^2+^]_e_-suppressible secretion dependent on the expression and signaling of homomeric CaSR. Both secretory modes are altered by aging to increase sPTH and sCa levels. We further linked VDR actions to GABA_B1_R/CaSR heterodimer-mediated tonic PTH secretion in PHPT patients. Close correlations between reduced VDR RNA levels with decreased expression of CASR RNA, increased expression of GABA_B1_R RNA, and increased GABA_B1_R/CaSR ratios (**fig. S2A, S2B**), suggested an increased likelihood of GABA_B1_R/CaSR heterodimerization at the expense of CaSR homodimers in parathyroid adenomas from PHPT patients compared to normal donor PTGs (see demographics of study subjects in **Table S1**). Additionally, upregulation of APP RNA expression correlated inversely with VDR RNA levels in PHPT adenomas versus control parathyroid glands, suggesting a role for APP in mediating PTH secretion (**fig. S2A, S2B**).

**Fig. 1.**
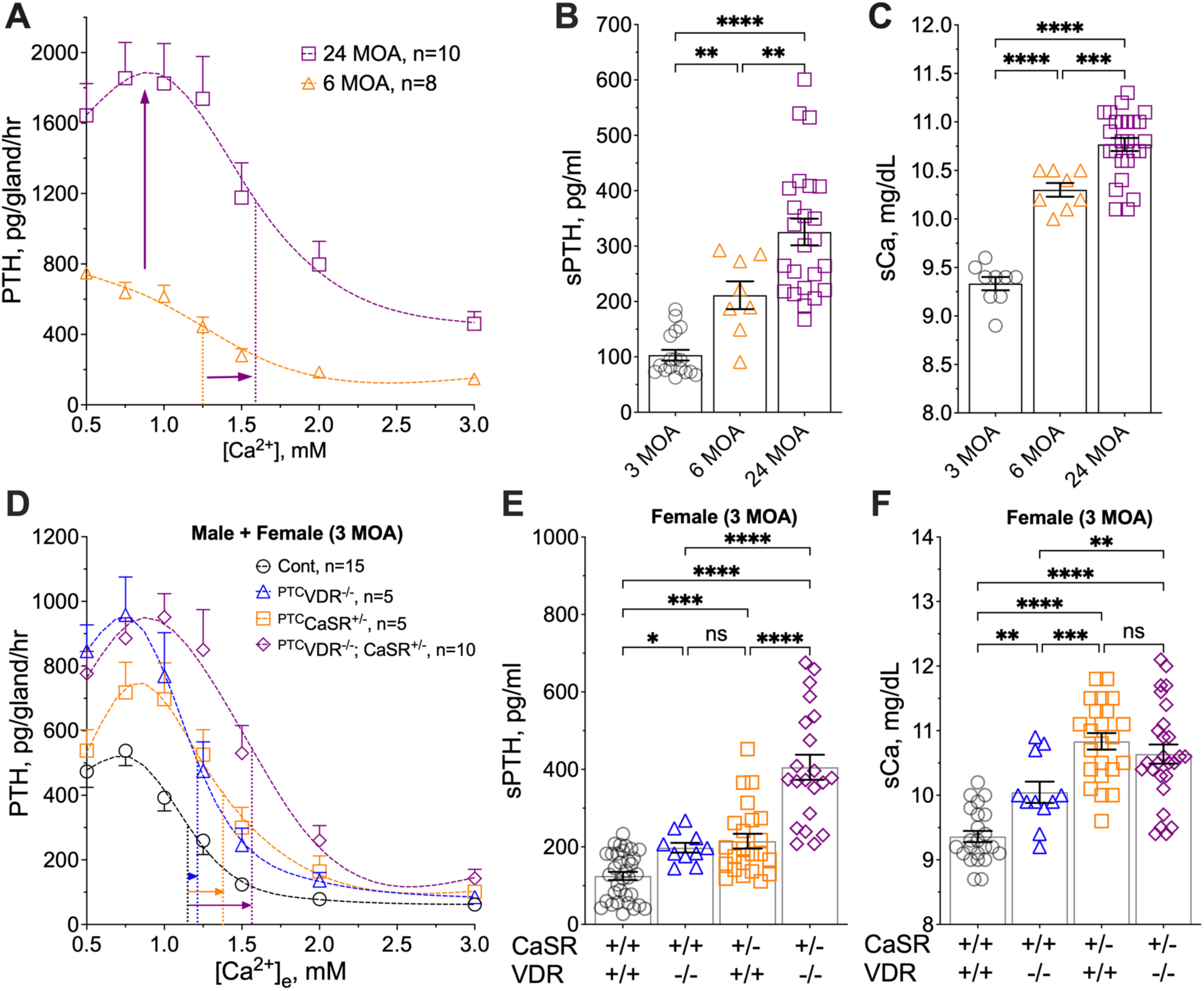
Distinct PTH secretory responses in aging and mouse models of HPT. (**A**) PTH/Ca^2+^ concentration-response curves were assessed in PTGs cultured from male C57/B6 mice at 6 (orange triangles) and 24 (purple squares) months of age (MOA) as previously described (*10*). Rates of PTH secretion (pg/gland/hour) at each [Ca^2+^]_e_ were plotted to extrapolate the maximal PTH secretion rate (PTH_Max_) and the Ca^2+^-setpoint (dotted lines). Mean ± s.e.m., *n*=8-10 mice per group. (**B**) Serum PTH (sPTH) and (**C**) total serum Ca^2+^ (sCa) of C57/B6 mice at 3 (black circles), 6 (orange triangles), and 24 (purple squares) MOA. Mean ± s.e.m., *n*=8-17 mice. (**D**) PTH/Ca^2+^ dose-response curves of PTGs from ^PTC^*Vdr^-/-^* (blue triangles), ^PTC^*Casr^+/-^* (orange squares), ^PTC^*Vdr^-/-^Casr^+/-^* (purple diamonds) mice and control littermates (black circles) at 3 MOA. Mean ± s.e.m., *n*=5-15 mice per genotype. (**E**) sPTH and (**F**) sCa of female ^PTC^*Vdr^-/-^*(blue triangles), ^PTC^*Casr^+/-^* (orange squares), ^PTC^*Vdr^-/-^Casr^+/-^* (purple diamonds) mice and control littermates (black circles) at 3 MOA. Mean ± s.e.m., *n*=10-32 mice. \**p*<0.05, \*\**p*<0.01, \*\*\**p*<0.005, \*\*\*\**p*<0.0001, and no significance (ns) was determined by one-way ANOVA with Fisher’s LSD test.

### APP and Aβ_42_ stimulate tonic PTH secretion in normal parathyroid tissue and PHPT adenomas

To determine whether APP and its cleavage products are altered in PHPT, we performed immunohistochemistry (IHC) and parathyroid-specific proteomic profiling assays (*20*) comparing the expression of key molecules in β-amyloidogenesis in PHPT adenomas versus normal parathyroid glands. The protein abundance of APP and its derivative peptide Aβ_42_ increased significantly in parathyroid adenomas compared to normal parathyroid tissue (**Fig. 2A, 2B**). Expression of BACE1 (β-secretase 1) and PSEN1 (presenilin 1, a subunit of γ-secretase) (**Fig. 2B**), was increased in PHPT adenomas, indicative of enhanced amyloidogenic processing of APP to produce Aβ_42_.

**Fig. 2.**
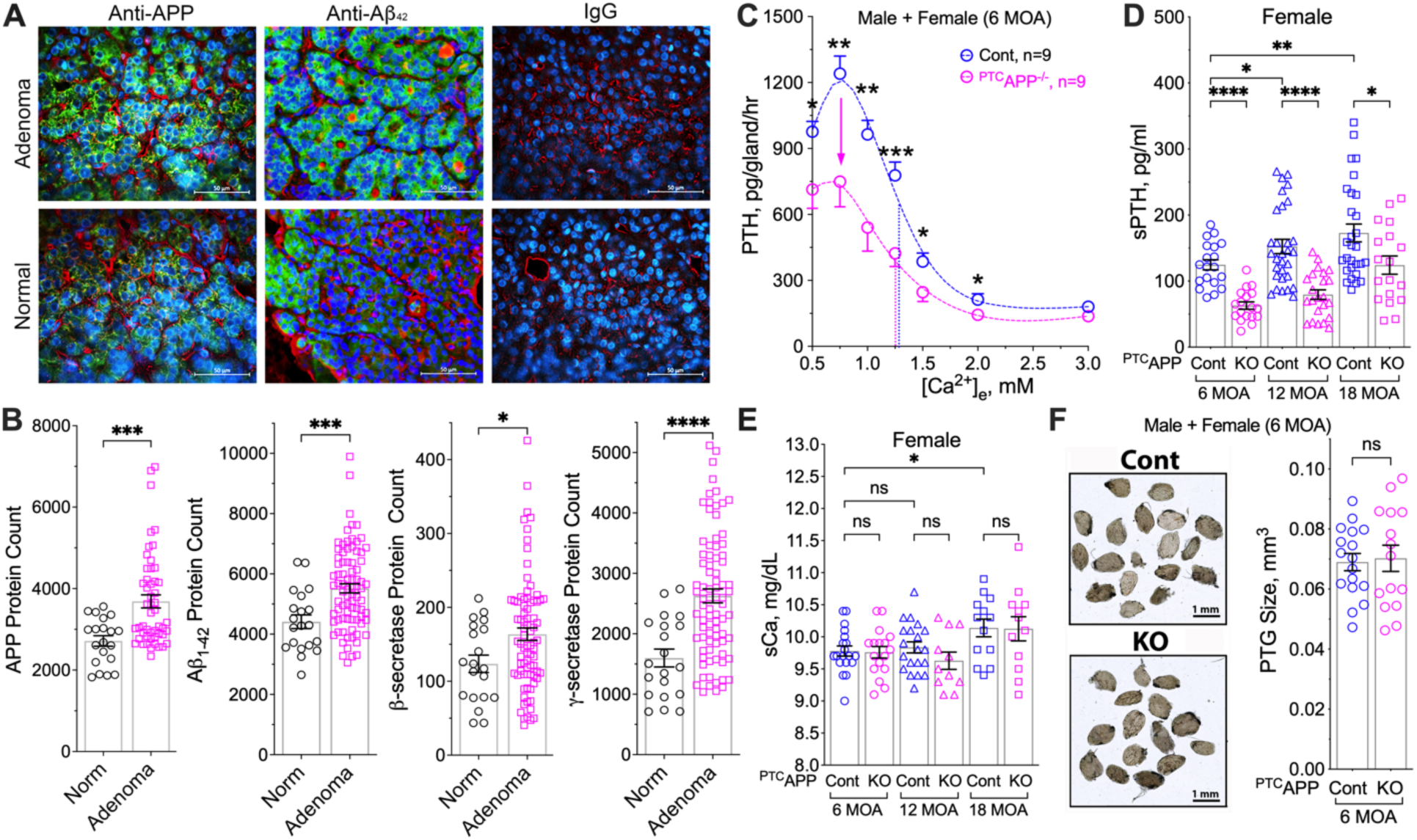
Expression of APP and Aβ_42_ in PTGs and the impact of *App* gene ablation on PTH secretory responses. (**A**) APP and Aβ_42_ (green) in human PTGs from normal donors (*n*=7) and in parathyroid adenomas from PHPT patients (*n*=8), counterstained with wheat germ agglutinin (red) for cell membrane and DAPI (blue) for nuclei. scale bar: 500 µm. (**B**) Proteomic profiling showed increased expression of APP, Aβ_42_, β-secretase, and γ-secretase in human parathyroid adenomas versus normal PTGs. Mean ± s.e.m., *n*=8-12. \**p* <0.05, \*\*\**p*<0.005, or \*\*\*\**p*<0.0001 (two-tailed student t-test). (**C**) PTH/Ca^2+^ dose-response curves of PTGs from *^PTC^App^-/-^*(pink circles) and *App*^fl/fl^ control (blue circles) at 6 MOA. Dotted vertical lines indicate Ca^2+^-setpoints for the corresponding genotypes. Mean ± s.e.m., of *n*=9 mice per genotype. (**D**) sPTH and (**E**) sCa levels of female *^PTC^App^-/-^* (pink symbols) and *App*^fl/fl^ control (blue symbols) mice at 6 (circle), 12 (triangle), and 18 (square) MOA. (**F**) Glandular volumes are equivalent between *^PTC^App^-/-^*(pink circles) and *App*^fl/fl^ control (blue circles). Scale bar: 1 mm, mean ± s.e.m. *n*=11-23 mice per group. ns (*p*>0.05), \**p*<0.05, \*\**p*<0.01, \*\*\*\**p*<0.0001 by one-way ANOVA with Fisher’s LSD test.

To determine whether APP and its cleavage products can causally modulate tonic PTH secretion, we compared rates of PTH secretion in parathyroid-specific *App* KO (^PTC^*App*^βflox/βflox^ or ^PTC^*App*^-/-^) mice to those in *App*^flox/flox^ control littermates. PTH_Max_ secretion was reduced by 33% in parathyroid glands from 6 MOA ^PTC^*App*^-/-^ mice versus control littermates, with no significant alteration in the Ca^2+^-setpoint (**Fig. 2C**). As anticipated, due to reduced tonic PTH secretion, baseline sPTH levels markedly decreased in both female (**Fig. 2D**) and male (**fig. S3A**) ^PTC^*App*^-/-^ mice relative to age-matched control littermates from 6 to 18 MOA. The reduction in sPTH was not due to alterations in sCa (**Fig. 2E** and **fig. S3B**) or differences in PTG sizes between ^PTC^*App*^-/-^ and control mice (**Fig. 2F**). These results indicate a non-redundant role for APP in sustaining ≈30% of tonic PTH secretion *in vivo*.

### Aβ_42_ enhances tonic PTH secretion via interactions with GABA_B1_R/CaSR heterodimers

We then compared the effects of two APP-derived peptides, Aβ_42_ and sAPP, on ex vivo PTH secretion, using parathyroid glands from ^PTC^*App*^-/-^ mice to avoid interference from endogenous APP. Aβ_42_ dose-dependently increased PTH secretion rate under normocalcemic conditions (**Fig. 3A**), while the 17-amino acid extension domain (ExtD, amino acids 204-220) peptide of sAPP-α and sAPP-β, which binds and activates GABA_B1_R in neurons (*11*), only modestly increased PTH secretion (**Fig. 3A**). A control peptide with the reverse ExtD sequence (amino acids 220-204) had no effect (**Fig. 3A**). Since Aβ_42_ (EC_50_=5.6 nM) is 10-fold more potent than the ExtD peptide (EC_50_=54.6 nM), our subsequent studies focused on the actions of Aβ_42_. Indeed, adding 1 μM of Aβ_42_ to ^PTC^*App*^-/-^ parathyroid glands effectively increased PTH_Max_ without shifting the Ca^2+^- setpoint when compared to vehicle or a reverse Aβ peptide (Aβ_42_-Rev) control (**Fig. 3B**). Similarly, Aβ_42_ (200 nM), but not the Aβ_42_-Rev peptide, elevated PTH_Max_ in human parathyroid adenoma tissue (**fig. S4**); the stimulatory effect could be blocked by a recombinant Aβ_42_- neutralizing antibody (**fig. S4**). The observed action of Aβ_42_ was likely mediated through GABA_B1_R, because addition of a competitive GABA_B1_R antagonist, CGP54626 (10 μM), blunted Aβ_42_-elicited PTH secretion in ^PTC^*App*^-/-^ parathyroid glands (**Fig. 3C**) and in normal and PHPT human parathyroid tissue (**Fig. 3D**).

**Fig. 3.**
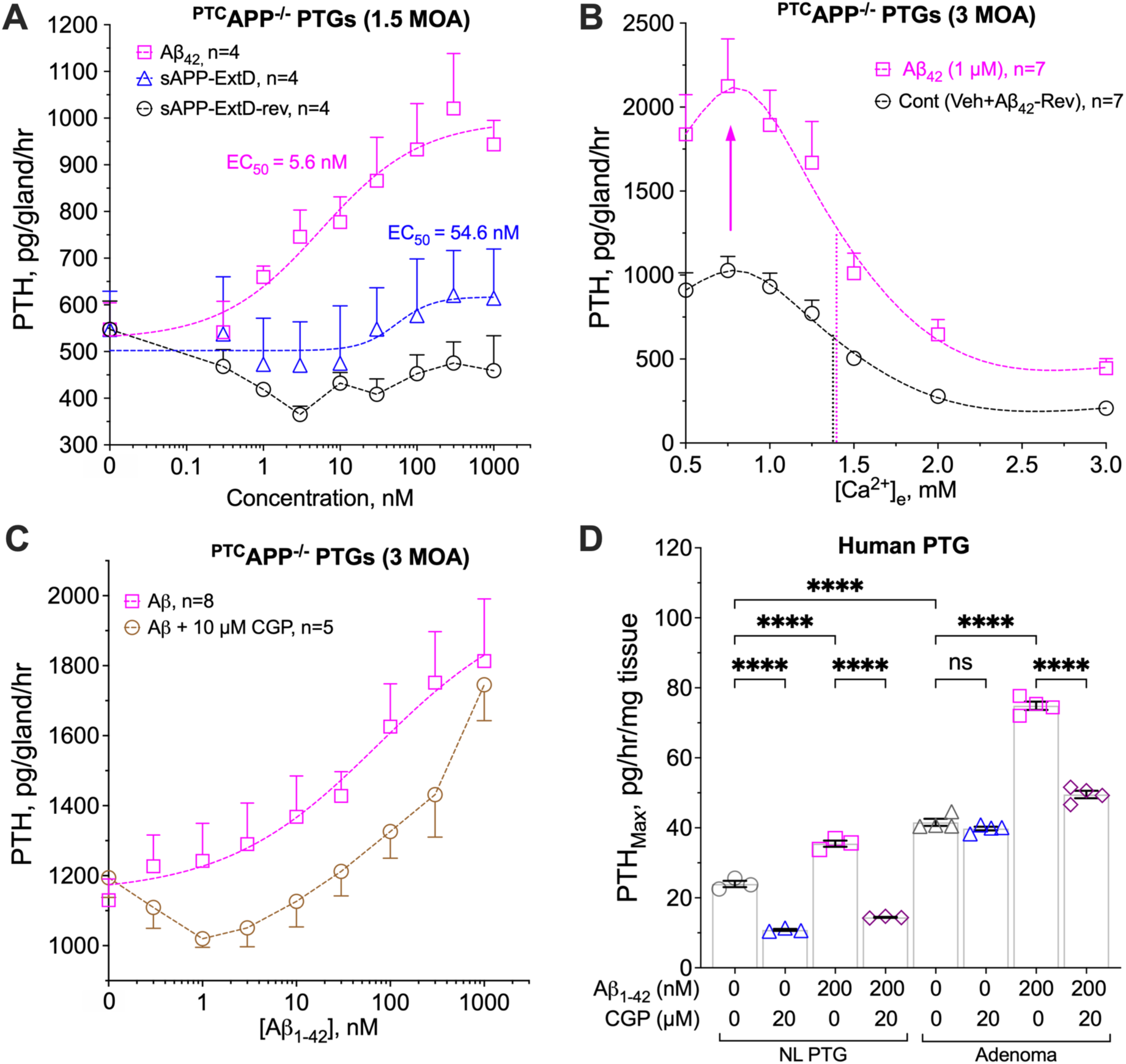
β-Amyloid enhances tonic PTH secretion. (**A**) PTGs of *^PTC^App^-/-^* mice were sequentially incubated with increasing concentrations (from 0. 3 to 1000 nM) of Aβ_42_ (pink squares), sAPP ExtD domain (sAPP-ExtD, blue triangles), or a reverse ExtD peptide (sAPP-ExtD-rev, black circles) in the presence of 1 mM [Ca^2+^]_e_ to determine changes in PTH secretion. (**B**) PTH/Ca^2+^ dose-response curves of PTGs from *^PTC^App^-/-^* mice in the presence of 1 µM of Aβ_42_ (pink squares), a reverse Aβ_42_ peptide (Aβ_42_-Rev), or vehicle (Veh, 0.1% DMSO). Aβ_42_-Rev and vehicle treatments were combined as the control group (black circles). Dotted vertical lines indicate Ca^2+^- setpoints for the corresponding treatments. (**C**) PTGs of *^PTC^App^-/-^* mice were sequentially incubated with increasing doses of Aβ_42_ with (brown circles) or without (pink squares) CGP54626 (CGP, 10 μM). Mean ± s.e.m., *n=*4-8 mice per treatment. (**D**) PTH_Max_ on a per mg of tissue and per hour basis was assessed in normal (NL) human PTGs and parathyroid adenomas from PHPT patients incubated with increasing [Ca^2+^]_e_ in the presence of Aβ_42_ peptide (200 nM) with or without CGP54626 (CGP, 20 μM). Mean ± s.e.m. of *n*=3-4 glands per treatment. \*\*\*\**p*<0.001 by two-way ANOVA with Sidak’s test.

Given the inability of the GABA_B1_R alone to localize to the cell surface and activate downstream G protein-mediated signaling (*21*) and the ability of CaSR to chaperone GABA_B1_R to the cell membrane of HEK-293 cells expressing both receptors (*22*), we proposed a direct action of Aβ_42_ on GABA_B1_R/CaSR heterodimers to initiate signaling responses that promote tonic PTH secretion. To test this hypothesis, we assessed the Aβ_42_-elicited PTH secretory responses in PTGs cultured from mice with parathyroid-specific deletion of *Gabbr1* and/or *Casr* in the background of ^PTC^*App*^-/-^. Indeed, the ability of Aβ_42_ to promote PTH secretion in the PTGs of ^PTC^*App*^-/-^ mice was completely abolished when *Gabbr1* (^PTC^*App*^-/-^*Gabbr1*^-/-^) and *Casr* (^PTC^*App*^-/-^*Casr*^-/-^) genes were ablated individually or concurrently (^PTC^*App*^-/-^*Gabbr1*^-/-^*Casr*^-/-^) (**Fig. 4A**). These data confirm the requirement for both GABA_B1_R and CaSR, presumably in the form of heterodimers, to mediate the PTH hypersecretory action of Aβ_42_. This notion was further supported by the ability of Aβ_42_ to instigate signaling responses of the GABA_B1_R/CaSR heterodimer that promote cAMP production (*10*). Specifically, we examined the impact of Aβ_42_ on homomeric CaSR-mediated G_i_ signaling by studying the high [Ca^2+^]_e_-induced inhibition of cAMP production in rat parathyroid gland-derived PTH-C1 cells pretreated with cholera toxin (CTx), which induces persistent activation of the Gs/adenylate cyclase/cAMP pathway. In CTx-treated PTH-C1 cells co-expressing CaSR and GABA_B1_R, activation of CaSR by high [Ca^2+^]_e_ (3 mM) promptly decreased accrual of cAMP, which was markedly lessened by subsequent administration of Aβ_42_ (1 μM) (**Fig. 4B**). This result indicates that suppression of G_i_ signaling and/or enhancement of G_s_ by Aβ_42_ occur through the activation of the GABA_B1_R/CaSR heterodimer to oppose CaSR homodimer signaling. Furthermore, we detected markedly increased colocalization (**Fig. 4C and fig. S5,** overlay, in yellow) of endogenous Aβ_42_ (in green) and GABA_B1_R/CaSR heterodimers (in red), stained by IHC and the proximity ligation assay (PLA), in parathyroid adenomas from PHPT patients with pre-operative vitamin D deficiency compared to normal donors or PHPT patients with replete 25OHD status. Collectively, these results strongly support the ability of Aβ_42_ to promote PTH hypersecretion through interactions with GABA_B1_R/CaSR heterodimers that antagonize Gi activation mediated by homomeric CaSR and perhaps other unknown pathways.

**Fig. 4.**
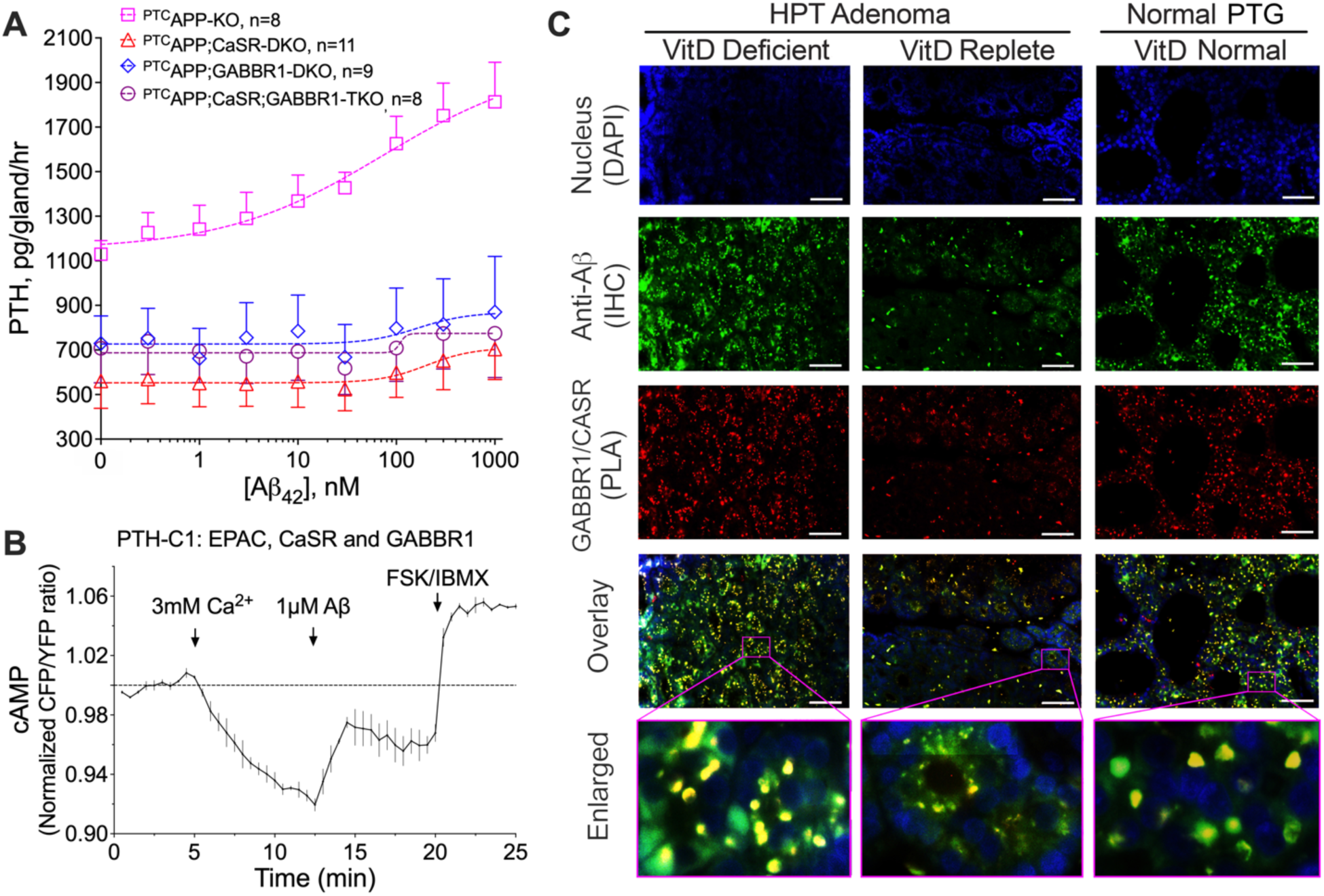
Aβ_42_ interacts with GABA_B1_R/CaSR heterodimers and antagonizes Gi activation. (**A**) PTGs from *^PTC^App^-/-^* (pink squares), *^PTC^App^-/-^Casr^-/-^*(red triangles), *^PTC^App^-/-^Gabbr1^-/-^* (blue diamonds), and *^PTC^App^-/-^Casr^-/-^Gabbr1^-/-^*(purple circles) mice were sequentially incubated with increasing doses (from 0. 3 to 1000 nM) of Aβ_42_ in the presence of 1 mM of [Ca^2+^]_e_. mean ± s.e.m., *n*=8-11 mice per genotype. (**B**) Averaged time-course of cAMP levels in the PTH-C1 cells co-expressed CaSR, GABA_B1_R, and a FRET-based cAMP reporter and were pretreated with cholera toxin. Arrows indicate the times to apply Ca^2+^ (3 mM), Aβ_42_ (1 μM), and forskolin/IBMX (10 μM), mean ± s.e.m. of *n*=3 experiments with 10–15 cells/experiment. (**C**) Detection of endogenous GABA_B1_R/CaSR heterodimers by the proximity ligation assay (PLA) and Aβ_42_ by immunohistochemistry (IHC) and their colocalization in overlayed images of human PTGs from normal donors and parathyroid adenomas from PHPT patients with deficient (<20 ng/ml) or repleted (>30 ng/ml) pre-operative 25OH vitamin D levels (*n*=4 glands per group). Aβ_42_ immunoreactivity was visualized with Alex Fluor 488 (green) and GABA_B1_R/CaSR with Texas Red (red) signals and counterstained with DAPI (blue) for nuclei. Colocalization of Aβ_42_ and GABA_B1_R/CaSR heterodimers was indicated by yellow signals in overlayed images. Scale bar: 125 µm.

### Vitamin D deficiency promotes PTH hypersecretion by upregulating APP/Aβ signaling responses

To quantitate the impact of vitamin D deficiency on signaling cascades of APP/Aβ and the GABA_B1_R/CaSR heterodimer, we compared parathyroid cell-defined proteomic profiles in the parathyroid tissue of normal donors and in adenomas from PHPT patients with deficient (<20 ng/ml) or replete (>30 ng/ml) pre-operative 25OH vitamin D levels. In line with the results from the ensemble RNA profiles (**fig. S2**), the proteomic profiles showed downregulation of CaSR and upregulation of GABA_B1_R in parathyroid tumors from vitamin D-deficient patients compared to vitamin D-replete patients and to normal controls, leading to a sharp elevation in the GABA_B1_R/CaSR ratio (**Fig. 5A and fig. S6**) predictive of an increased propensity for GABA_B1_R/CaSR heterodimerization. Expression of APP and Aβ_42_ were also markedly increased in parathyroid tumors from vitamin D-deficient PHPT patients; however, these changes were significantly mitigated in patients with replete 25OHD status (**Fig. 5B and fig. S6**). Likely due to the ability of Aβ_42_ accrual to induce accumulation and hyperphosphorylation of Tau (*23*), the abundance of Tau protein and its phosphorylation at serine-404 and serine-396 was markedly increased in tumors from vitamin D-deficient PHPT patients. These effects were completely reversed in the tumors from PHPT patients with replete pre-operative vitamin D status (**Fig. 5C and fig. S6**). These findings support a role for Tau as a downstream effector of Aβ_42_ in promoting PTH hypersecretion in the context of vitamin D deficiency. Indeed, blocking Tau phosphorylation with the potent protein kinase inhibitor K252a (*24*) effectively eliminated the ability of Aβ_42_ to stimulate PTH secretion in mouse parathyroid glands (**Fig. 5D**).

**Fig. 5.**
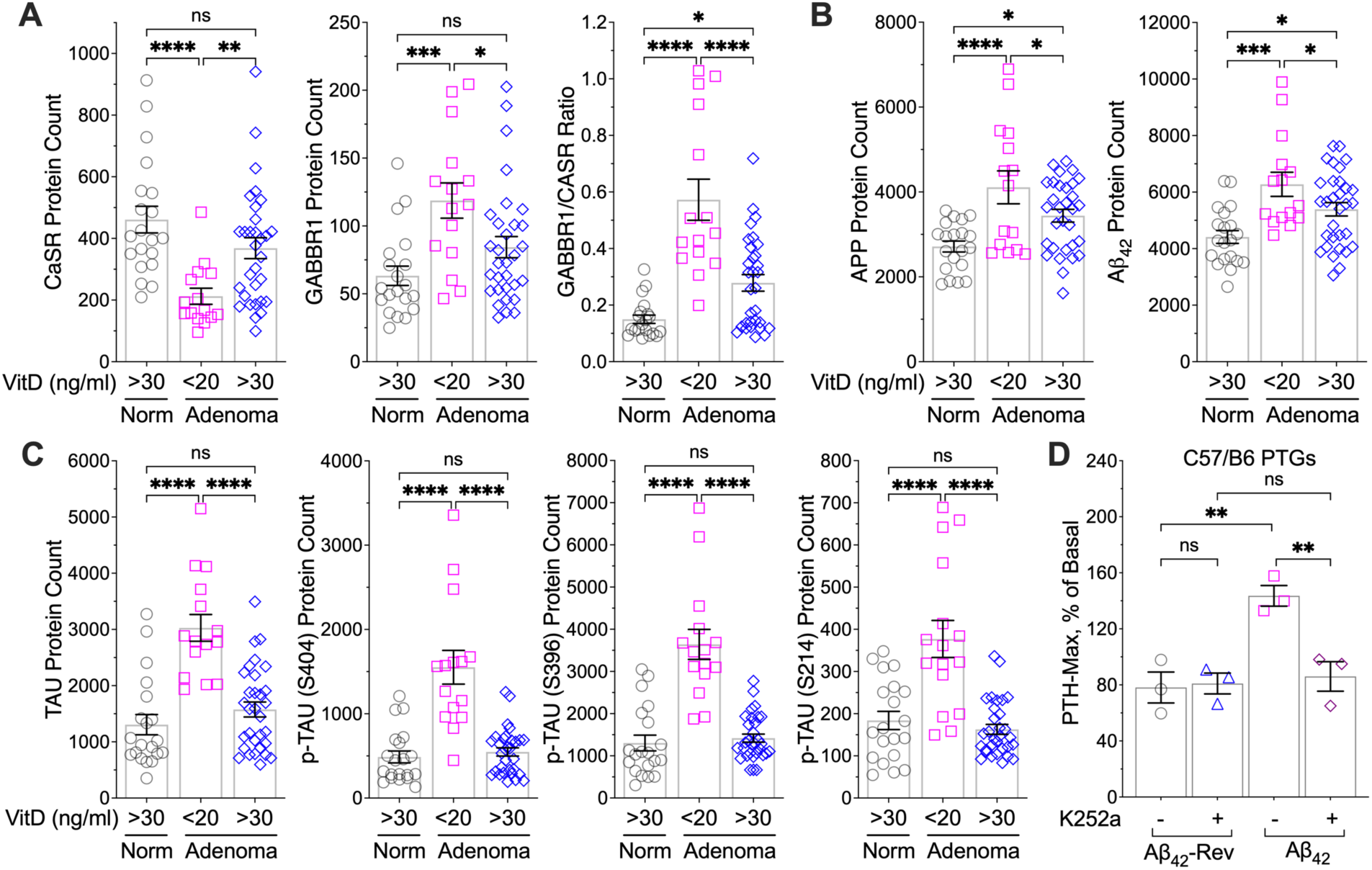
Impact of vitamin D deficiency on APP/Aβ signaling responses. Protein levels of (**A**) CaSR and GABA_B1_R, (**B**) APP and Aβ_42_, and (**C**) Tau and its phosphorylation at Serine 404, 396, and 214 were assessed by spatial proteomic profiling of human PTGs of normal donors (norm, *n*=7, grey circles) and parathyroid adenomas of PHPT patients with vitamin D deficiency (pre-operative 25OHD <20 ng/ml, *n*=8, pink squares) or replete vitamin D status (25OHD≥30 ng/ml, *n*=15, blue diamonds). Mean ± s.e.m. (**D**) PTGs of 6-week-old wild-type C57/B6 mice were sequentially incubated with increasing doses (from 0. 3 to 1000 nM) of Aβ_42_ or a control peptide with reverse sequence (Aβ_42_-Rev) with or without protein kinase inhibitor K252a (1 μM) in the presence of 1 mM of [Ca^2+^]_e_. Changes in the rate of PTH secretion on a per gland and per hour basis were assessed, and the maximal PTH secretion rate (PTH_Max_) was presented after normalized to the basal rate prior to treatment. Mean ± s.e.m. of *n*=3 mice per treatment. ns (*p*>0.05), \**p*<0.05, \*\**p* <0.01, \*\*\**p*<0.005, and \*\*\*\**p*<0.0001 by one-way ANOVA with Fisher’s LSD test.

To confirm the role of APP in driving PTH hypersecretion amid vitamin D deficiency, we compared the secretory responses of parathyroid glands from ^PTC^*Vdr*^-/-^ mice with or without concurrent parathyroid-specific ablation of the *App* gene (^PTC^*Vdr*^-/-^*App*^-/-^). Blocking VDR expression significantly increased PTH_Max_ with a small shift of the Ca^2+^-setpoint to the right (**Fig. 1D and Fig. 6A**). Because of the increased tonic PTH secretion and Ca^2+^-set-point, the sPTH and sCa levels in 12-month-old ^PTC^*Vdr*^-/-^ mice were significantly higher than those in control littermates (**Fig. 6B, 6C and fig. S7**). Concurrent ablation of the *App* gene in the ^PTC^*Vdr*^-/-^*App*^-/-^ mice abrogated the PTH hypersecretion seen in ^PTC^*Vdr*^-/-^ mice but only partially restored the Ca^2+^-setpoint (**Fig. 6A**), indicating that the VDR mediates both APP-dependent and APP-independent actions in regulating PTH secretion. Nonetheless, serum PTH levels in the ^PTC^*Vdr*^-/-^*App*^-/-^ mice were restored to the level of control littermates along with normalized serum Ca^2+^ levels (**Fig. 6B, 6C and fig. S7A, S7B**). Supporting a role for Tau in mediating Aβ_42_-induced tonic PTH secretion, parathyroid glands cultured from *Mapt*^-/-^ mice, in which the Tau-encoding *Mapt* gene is globally deleted, displayed a reduction in PTH_Max_ levels relative to control littermates (**Fig. 6D**). Concurrent ablation of *Mapt* and *Vdr* in ^PTC^*Vdr*^-/-^//*Mapt*^-/-^ double KO mice prevented the stimulatory effects of *Vdr* KO on tonic PTH secretion (**Fig. 6D**). These results support a signaling relationship where Tau is an essential downstream effector of APP/Aβ in promoting tonic PTH secretion in the context of vitamin D deficiency.

**Fig. 6.**
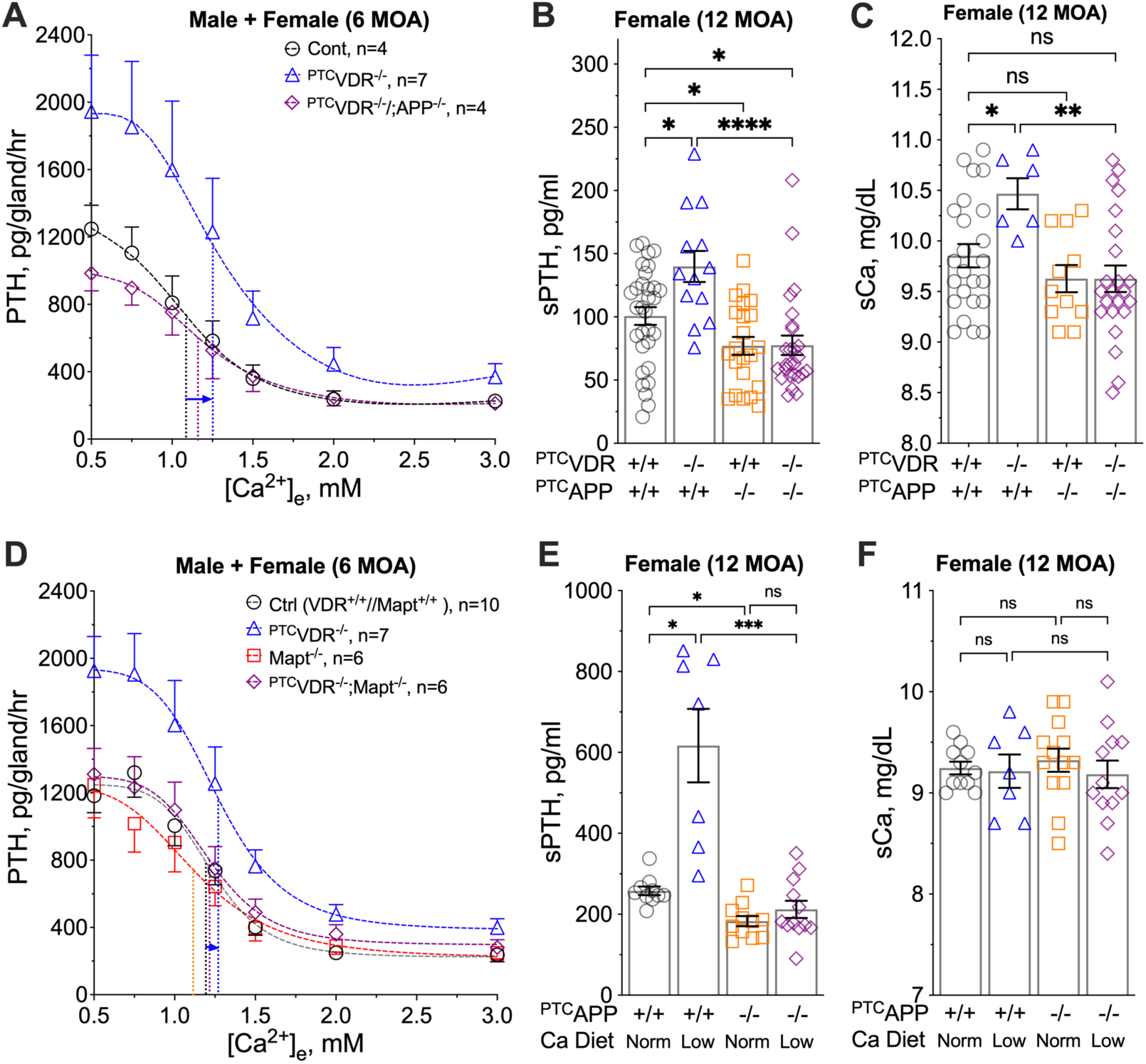
Ablation of the parathyroid *App* gene prevented PTH hypersecretion in mouse models of vitamin D deficiency and chronic Ca^2+^ deficiency. (**A**) PTH/Ca^2+^ dose-response curves of PTGs from ^PTC^*Vdr*^-/-^ (blue triangles), ^PTC^*Vdr*^-/-^*App*^-/-^ (purple diamonds), and control littermates (grey circles) at 6 MOA. Dotted vertical lines indicate the Ca^2+^-setpoints. mean ± s.e.m., of *n*=4-7 mice per genotype. (**B**) sPTH and (**C**) sCa of female ^PTC^*Vdr*^-/-^ (blue triangles), ^PTC^*App*^-/-^ (orange squares), ^PTC^*Vdr*^-/-^*App*^-/-^ (purple diamonds) mice, and controls (*Vdr*^+/+^*App*^+/+^, grey circles) at 12 MOA. mean ± s.e.m., *n*=6-23 mice per genotype. ns (*p*>0.05), \**p*<0.05, \*\**p*<0.01, \*\*\*\**p*<0.0001 by one-way ANOVA with Fisher’s LSD test. (**D**) PTH/Ca^2+^ dose-response curves of PTGs from ^PTC^*Vdr*^-/-^ (blue triangles), *Mapt*^-/-^ (orange circles), ^PTC^*Vdr*^-/-^//*Mapt*^-/-^ (purple diamonds) mice and control littermates (*Vdr*^+/+^*Mapt*^+/+^, grey circles) at 6 MOA. mean ± s.e.m., of *n*=6-10 mice per genotype. (**E**) sPTH and (**F**) sCa levels of male ^PTC^*App*^-/-^ and *App*^fl/fl^ control littermates at 12 MOA after feeding with normal (1%) or low calcium (0.02%) diets for 6 weeks. Mean ± s.e.m. of *n*=6– 12 mice for each group, \**p*<0.05 and \*\**p*<0.01 between groups indicated by one-way ANOVA with Sidak’s test.

To test whether the APP/Aβ pathway is also involved in PTH hypersecretion secondary to chronic Ca^2+^-deficiency, we subjected ^PTC^*App*^-/-^ mice and their control littermates to a continuous low Ca^2+^ diet (0.02% vs 1% in the normal diet) for 6 weeks. The increase of serum PTH levels (by ≈50%) induced by low Ca^2+^ diet in control littermates was completely absent in the ^PTC^*App*^-/-^ mice (**Fig. 6E**), despite comparable serum Ca^2+^ levels (**Fig. 6F**). This result is consistent with a role for the APP/Aβ pathway in mediating PTH hypersecretion in secondary hyperparathyroidism.

### An Aβ-neutralizing antibody suppresses tonic PTH secretion and reduces serum PTH levels

Serum PTH levels rise in the elderly due to multiple contributing factors (*25, 26*). We observed an upregulation of Aβ_42_ expression with age from 6 to 18 MOA in the parathyroid glands of male *App*^fl/fl^ control, but not ^PTC^*App*^-/-^, mice (**fig. S8**), suggesting a role for the Aβ signaling cascade in promoting PTH hypersecretion in aging mice. We tested whether reducing Aβ_42_ levels using the recombinant Aβ-neutralizing antibody aducanumab (Adu), developed to reduce Aβ_42_ loads and slow cognitive decline in Alzheimer’s Disease (AD) patients (*19*), could lessen aging-associated hyperparathyroidism. We first compared PTH secretion in response to increasing [Ca^2+^]_e_ in parathyroid glands from mice treated with murinized aducanumab (*19*), an isotype-matched IgG, or a calcimimetic agent (cinacalcet) that activates CaSR homodimers. Parathyroid glands from 24-month-old male C57/B6 mice treated with the control IgG exhibited PTH hypersecretion, with a significant increase in PTH_max_ and right-shifted Ca^2+^-setpoint (**Fig. 7A**) compared to younger (6 MOA) mice (**Fig. 1A**). Ex vivo incubation of 24-month-old parathyroid glands with aducanumab (50 µg/ml) significantly reduced PTH secretion rates without affecting the Ca^2+^ setpoint (**Fig. 7A**). In contrast, cinacalcet (50 nM) markedly reduced both PTH_max_ and the Ca^2+^ setpoint (**Fig. 7A**), consistent with its actions as a positive allosteric modulator (PAM) of CaSR homodimers (*27*). *In vivo* administration of aducanumab alleviated hyperparathyroidism symptoms in aged mice. Low (10 mg/kg/week by once weekly injection for 5 weeks) and high (40 mg/kg/week by twice weekly injections for 5 weeks) doses of aducanumab reduced sPTH to a similar degree (**Fig. 7B**), suggesting that a significant pool of PTH (≈34% of total serum PTH) is mediated by Aβ_42_. In contrast, daily injections of cinacalcet (5 mg/kg, once daily) transiently reduced sPTH (**Fig. 7B**) to <30% of total sPTH but produced much more severe hypocalcemia (**Fig. 7C**). Intriguingly, co-treatment with aducanumab further reduced sPTH while partially normalizing cinacalcet-induced hypocalcemia (**Fig. 7B and 7C**).

**Fig. 7.**
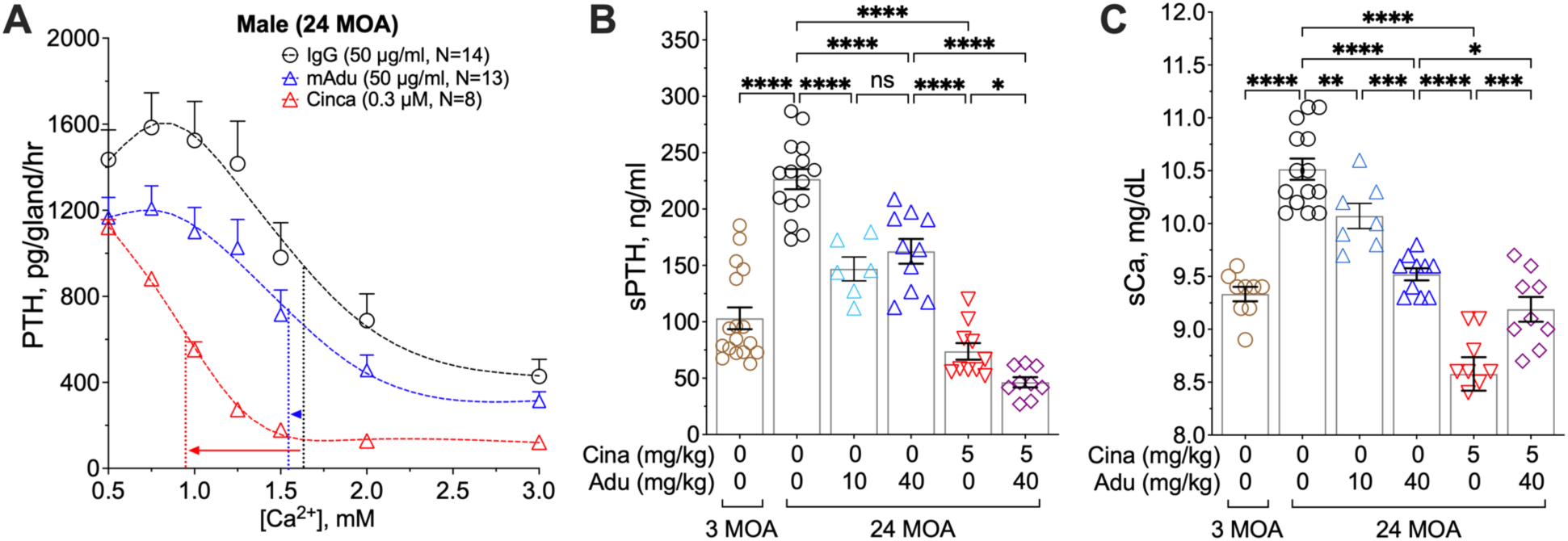
Neutralizing Aβ suppresses tonic PTH secretion and reduces serum PTH levels in HPT due to aging. (**A**) Changes in PTH secretion rate in PTGs from 24 month-of-old C57/B6 mice incubated with control murine IgG (50 µg/ml, black circles), cinacalcet (Cina, 50 nM, red triangles), or Aducanumab (mAdu, 50 µg/ml, blue triangles) were assessed in the presence of increasing [Ca^2+^]_e_. Dotted vertical lines indicate Ca^2+^-setpoints of the corresponding treatment. mean ± s.e.m., of *n*=6-15 mice per treatment. (**B**) sPTH and (**C**) sCa of male C57/B6 mice after twice-weekly injections of vehicle (brown circles: 3 MOA; black circles: 24 MOA), daily injections of cinacalcet (Cina, 5 mg/kg/day, red triangles), a low dose of aducanumab (Adu, 10 mg/kg/week by once weekly injection, cyan triangles) or a high dose (40 mg/kg/week by twice weekly of 20 mg/kg injections, blue triangles), or a combination of cinacalcet and aducanumab (purple diamonds). mean ± s.e.m., of *n*=7-15 mice for each treatment. ns (*p*>0.05), \*\**p*<0.01, \*\*\**p*<0.005, \*\*\*\**p*<0.0001 by one-way ANOVA with Fisher’s LSD test.

## DISCUSSION

Prior studies of parathyroid adenomas from PHPT patients revealed two distinct modes of PTH hypersecretion: one group displayed reduced Ca^2+^ sensitivity (higher Ca^2+^-setpoints) with lower levels of tonic PTH (PTH_Max_), while the other group retained a normal Ca^2+^-setpoint with greater tonic levels of PTH (*7*). These distinct secretory phenotypes were closely replicated in two PHPT mouse models. In a model of familial hypocalciuric hypercalcemia (*^PTC^Casr^+/-^* mice), deletion of one allele of *Casr* in parathyroid cells led to a profoundly elevated Ca^2+^-setpoint with a moderate increase in PTH_Max_. In contrast, *^PTC^Vdr^-/-^* mice (mimicking chronic vitamin D deficiency) had much higher PTH_Max_ with smaller impact on Ca^2+^-setpoint. These alternative modes of PTH hypersecretion revealed distinct actions of the CaSR and the VDR that may underlie the divergent secretory patterns seen in different types of parathyroid adenomas.

We have shown previously that heterodimerization with GABA_B1_R alters CaSR signaling to increase PTH_Max_ and Ca^2+^-setpoint of parathyroid cells (*10*). Blocking the production of endogenous GABA in parathyroid cells lowered the Ca^2+^-setpoint, but not PTH_Max_, leading to a modestly reduced sPTH level amid hypocalcemia (*10*). Our current work uncovers Aβ as an endogenous activator of GABA_B1_R/CaSR heterodimers, capable of increasing PTH_Max_ without altering the Ca^2+^-setpoint of parathyroid cells. Aβ can confer Ca^2+^-independent production of serum PTH, as demonstrated by the chronically reduced sPTH levels in ^PTC^*App*^-/-^ mice in the presence of normocalcemia. These findings reveal a novel autocrine/paracrine mechanism influencing PTH secretion beyond Ca^2+^ sensing and demonstrate unique ligand-dependent biased signaling of dimeric (or multimeric) family C GPCRs in altering cell functions. Furthermore, our observations of (i) a complete loss of Aβ_42_-mediated stimulatory effect on PTH secretion in parathyroid glands lacking GABA_B1_R or CaSR; (ii) colocalization of endogenous Aβ_42_ with GABA_B1_R/CaSR heterodimers in parathyroid cells; and (iii) the ability of Aβ_42_ to acutely diminish homomeric CaSR-mediated Gi activation in PTH-C1 cells co-expressing GABA_B1_R, collectively support a role for the GABA_B1_R/CaSR heterodimer as an Aβ_42_ receptor.

Multiple lines of evidence from our data support a causal link between vitamin D deficiency and enhanced Aβ signaling in parathyroid cells. First, the secretory profile of parathyroid glands from ^PTC^*Vdr*^-/-^ mice closely resembles the profile induced by exogenous Aβ_42_. Second, transcriptomic analyses of parathyroid adenomas show an inverse relationship between APP and VDR RNA expression. Third, proteomic analyses of PHPT adenomas shows enhanced β-amyloidogenesis in tumors from patients with pre-operative vitamin D deficiency but not in those from vitamin D replete patients. Lastly, depleting endogenous Aβ in parathyroid cells completely reverses the PTH hypersecretory output and hyperparathyroidism phenotype seen in ^PTC^*Vdr*^-/-^ mice. These studies establish a new and long-sought biochemical connection linking the action of VDR to PTH hypersecretion mediated *via* Aβ signaling.

Upregulated expression and phosphorylation (particularly, at serine-404 and serine-396) of Tau are closely correlated with increased β-amyloidogenesis in parathyroid adenomas from patients manifesting vitamin D deficiency. These changes were not seen in tumors from patients with replete vitamin D status, supporting a role of phospho-Tau as a downstream effector of Aβ_42_ signaling in parathyroid cells, specifically in the context of vitamin D deficiency. This notion is further supported by the inability of VDR KO to enhance tonic PTH secretion in the PTGs lacking the Tau-encoding *Mapt* gene (**Fig. 6D**). Prior studies of Alzheimer’s disease have shown that Aβ-induced Tau hyperphosphorylation at serine-396 and serine-404 is one of the earliest events responsible for the functional loss of Tau-mediated tubulin polymerization, triggering neurodegeneration (*28*). Aβ induces Tau hyperphosphorylation potentially by activating multiple protein kinases, including cAMP-dependent protein kinase (PKA), AMP-activated protein kinase (AMPK), GSK-3β, cyclin-dependent protein kinase 5 (cdk5), mitogen-activated protein kinase (MAPK), and the Src-family kinase (*29*). Supporting a role for increased Aβ-induced protein kinase activity in promoting PTH hypersecretion, suppression of Tau phosphorylation with K252a, a potent protein kinase inhibitor (*24*), effectively blocks the ability of Aβ_42_ to stimulate PTH secretion in both murine and human PTGs. Future studies will be required to identify the specific kinases that modify Tau in the parathyroid and to delineate their actions in coupling Aβ stimulation to Tau phosphorylation and PTH secretion.

Consolidating these results, we propose a conceptual model connecting vitamin D deficiency, APP/Aβ_42_ signaling, and PTH hypersecretion. Activation of the VDR by 1,25(OH)_2_D under normal conditions suppresses expression of APP and limits proteolytic processing towards Aβ_42_ production by inhibiting β- and γ-secretase expression. Concurrently, VDR signaling increases CaSR and suppresses GABA_B1_R expression in parathyroid cells, promoting homomeric CaSR-mediated G_q_ and G_i_ signaling and reducing the heterodimerization of GABA_B1_R and CaSR, thus preventing high levels of tonic PTH secretion (**Fig. 8A**). Conversely, aging and other factors that chronically reduce 1,25(OH)_2_D levels and/or parathyroid VDR expression attenuate the actions of VDR in parathyroid cells (**Fig. 8B**), resulting in increased GABA_B1_R expression and decreased CaSR levels, favoring GABA_B1_R/CaSR heterodimerization at the expense of CaSR/CaSR homodimers. Reduced VDR activity enhances the expression of the *APP, BACE1, and PSEN1* genes and increases the production of Aβ_42_, which directly or indirectly activates GABA_B1_R/CaSR heterodimers to increase cAMP production and enhance the expression and phosphorylation of Tau, leading to PTH hypersecretion and the manifestations of hyperparathyroidism.

**Fig. 8.**
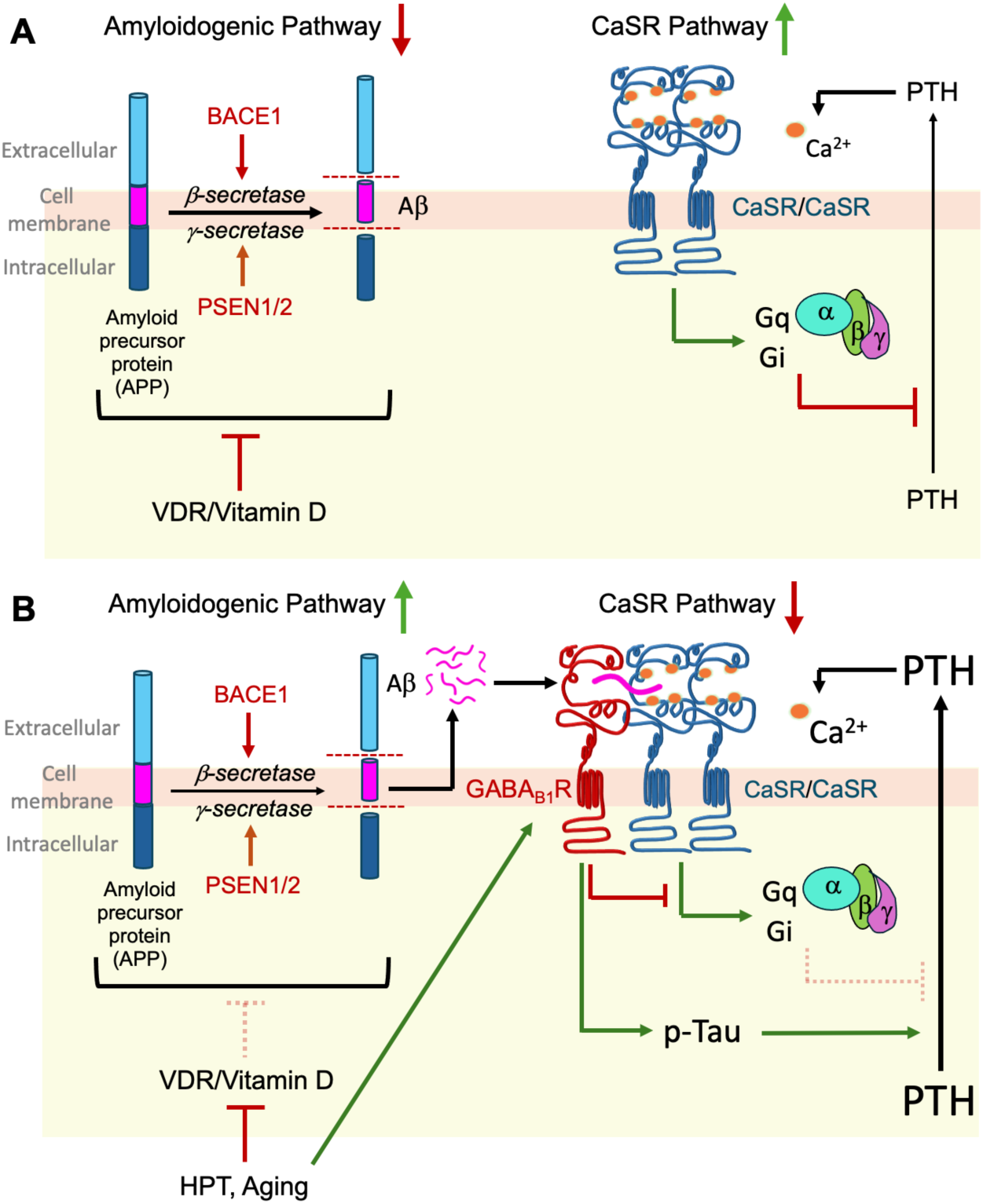
A working model for APP/Aβ_42_ signaling in promoting PTH hypersecretion in HPT secondary to vitamin D deficiency and aging. **(A)** Activation of VDR in parathyroid cells by 1,25(OH)_2_D restricts Aβ_42_ levels by suppressing the expression of APP and β- and γ-secretase. Activated VDR also increases CaSR expression and enhances the homomeric CaSR-mediated G_q_ and G_i_ signaling to limit tonic PTH secretion. **(B)** In aging and HPT states, VDR and CaSR activities in parathyroid cells decline due to chronic vitamin D deficiency, leading to increased Aβ_42_ and GABA_B1_R expression and the formation of GABA_B1_R/CaSR heterodimer. Activation of GABA_B1_R/CaSR heterodimers by Aβ_42_ antagonizes the G_q_ and G_i_ signaling and promotes phosphorylation of Tau, driving PTH hypersecretion.

While our current study focused on establishing a link between Aβ signaling and PHPT in the context of vitamin D deficiency, a fundamental role of Aβ in mediating PTH hypersecretion may extend to other forms of hyperparathyroidism. This notion is supported by the inability of a long-term (six weeks) low Ca^2+^-diet to elevate sPTH levels in ^PTC^*App*^-/-^ mice (**Fig. 6E**) and the ability of an Aβ-neutralizing antibody to reduce sPTH levels in aging mice (**Fig. 7B**). In addition, prior studies showed profound upregulation of the GABA_B1_R/CaSR heterodimer in hyperplastic PTGs from patients with secondary hyperparathyroidism (SHPT) (*10*), a clinical cohort where vitamin D deficiency is a significant contributor (*8*).

If future studies confirm that aberrantly enhanced Aβ signaling is a common driver of PTH hypersecretion, interfering with this pathway could form the basis of a new and actionable therapeutic approach for hyperparathyroidism, particularly in cases where surgical management is not indicated, appropriate, or safe (*27*). Our data in aging mice suggest several potential advantages for neutralizing Aβ signaling with aducanumab when compared to the conventional therapy replying on calcimimetic agents such as cinacalcet to activating homodimeric CaSRs. First, the longer half-life of aducanumab compared to cinacalcet could reduce administration frequency, following established dosing patterns recommended for aducanumab in the treatment of Alzheimer’s disease. Second, targeting the upregulated APP/Aβ signaling pathway with aducanumab has the favorable capacity to reduce PTH hypersecretion and normalize high sPTH in aged mice with a milder reduction in sCa levels compared to cinacalcet. Antagonism of Aβ_42_-mediated signaling in parathyroid cells may thus provide a beneficial, alternative Ca^2+^-sparing approach to treat SHPT patients at elevated risk for iatrogenic hypocalcemia and seizures. This effect of aducanumab is likely due to its ability to target solely the PTH pool responsible for increasing tonic PTH levels with minimal impact on the Ca^2+^-suppressible pools of PTH controlled by CaSR/CaSR homodimers, allowing parathyroid cells to continue to serve as a “calciostat” in maintaining Ca^2+^ homeostasis. Even at a saturation dose for suppressing the tonic pools of PTH (**Fig. 7B**), aducanumab reversed aging-induced hypercalcemia to serum calcium levels only slightly higher than those in 3-month-old vehicle-treated mice (**Fig. 7C**). In contrast, while cinacalcet was much more effective in transiently suppressing sPTH levels, it caused more severe hypocalcemia. Intriguingly, the hypocalcemic effect of cinacalcet could be partially mitigated by co-treatment with aducanumab (**Fig. 7C**), even though the two drugs synergistically suppressed sPTH (**Fig. 7B**). How the hypocalcemic effect of cinacalcet was reduced by aducanumab remains unclear. However, the interaction of these two agents suggests a potentially advantageous combination therapy, particularly in SHPT patients with chronic kidney disease, for whom calcium management can be particularly challenging. Third, aducanumab (*19*) and its comparable analog, lecanemab (*30*), have been approved by the US FDA to reduce Aβ loads in the CNS of patients with Alzheimer’s Disease. The extensive published safety data and side-effect studies supporting clinical implementation in AD could streamline repurposing Aβ-neutralizing biological agents for the treatment of hyperparathyroidism.

While the work presented here reveals a new, targetable signaling pathway that drives PTH hypersecretion associated with aging and vitamin D deficiency, important study limitations must be considered. The genetic data are fully consistent with an obligatory role for both CaSR and GABA_B1_R in mediating the hypersecretory response to Aβ_42_, but we cannot rule out the possibility of other biochemical targets for the peptide in the absence of direct binding studies and formal demonstration of Aβ_42_ interaction sites that uniquely regulate the activity of the CaSR/GABA_B1_R heterodimer. The increased abundance of phosphorylated Tau in parathyroid adenomas may be a consequence rather than a driver of hypersecretory behavior, especially considering the broad and relatively non-selective biochemical activity of the K562 kinase inhibitor. The observed capacity of the Aβ_42_-neutralizing antibody aducanumab to mitigate PTH hypersecretion *in vivo* in the murine models and *ex vivo* in human parathyroid tissue presents an intriguing possibility for repurposing an FDA-approved agent for medical management of hyperparathyroidism. To facilitate this drug repurposing, future studies will be required to confirm Aβ_42_ neutralization as the operative mechanism for the PTH suppressive response to aducanumab or other similar biologic agents, define the nature of Aβ_42_ suppressive interactions with calcimimetic compounds on PTH end organ targets, and conduct careful evaluation of the action of these compounds in other pathophysiological contexts such as chronic renal failure where additional tools for medical management of PTH hypersecretion would be especially useful. Developing strategies to exploit the novel biochemical pathway reported here could provide the basis for a much-needed alternative approach for managing hyperparathyroidism.

## MATERIALS AND METHODS

### Study design

The objectives of the study were to test the hypothesis that APP and its derivative β-Amyloid drive tonic PTH hypersecretion via interactions with GABA_B1_R/CaSR heterodimers. We evaluated the impact of APP/Aβ_42_ on parathyroid secretory properties using *ex vivo* PTG cultures from human donors and various genetically manipulated mice and investigated its regulation by vitamin D with transcriptomic and proteomic profiling in parathyroid tissues from patients with primary HPT and normal donors.

De-identified normal human PTGs were obtained through our institution’s solid organ transplant service from an unselected sequential series of eucalcemic donors (*n* = 7), using a fully authorized tissue procurement protocol for the recovery of viable, intact PTGs. Donor glands were confirmed as histologically normal by a board-certified attending pathologist with expertise in parathyroid histopathology. Parathyroid adenoma specimens from patients undergoing surgery for PHPT (*n* = 40) were obtained with informed consent. Clinical, demographic, pathological, and histopathological patient data were collected from the medical record and anonymized by the study clinical research coordinator in compliance with IRB requirements (table S1). All patient identifiers were deleted, and samples were stored and processed with unique numerical identifiers. PHPT patients met the following clinical criteria: (i) serum Ca ≥ 10.2 and ≤ 13.5 mg/dL; (ii) elevated serum PTH levels in setting of hypercalcemia; and (iii) the absence of a known genetic basis for the PHPT (e.g., MEN1). All human tissue collection and study protocols were approved by the University of California-San Francisco Institutional Review Board (IRB protocol number 19-27072). For PLA assays, immunohistochemical staining, and spatial proteomic profiling, PTGs were fixed in 4% PFA, embedded in paraffin, and sectioned at 5-mm thickness before processing as described below.

Several genetically manipulated mouse lines were used in this study. Wild-type C57/B6, PTH-Cre, *App^flox/flox^* (RRID:IMSR_JAX:030770), and global *Mapt*^-/-^-null mice (RRID:IMSR_JAX:007251) were purchased from Jackson Laboratory (JAX; Bar Harbor, Maine, USA). *Vdr^flox/flox^* mice were provided by Dr. Shigeaki Kato (University of Tokyo, Japan) (*31*). *Gabbr1^flox/flox^* mice were provided by Dr. Bernhard Bettler (University of Basel, Switzerland) (*32*). *Casr^flox/flox^*mice were made previously (*33*). PTH-Cre, *App^flox/flox^*, *Gabbr1^flox/flox^*, and *Casr^flox/flox^* mice were bred to generate *^PTC^App^-/-^, ^PTC^App^-/-^Gabbr1^-/-^*DKO, and *^PTC^App^-/-^Casr^-/-^* DKO mice, which were then used to produce *^PTC^App^-/-^Gabbr1^-/-^Casr^-/-^*triple KO mice. Control littermates carry *App^flox/flox^*, *App^flox/flox^Gabbr1^flox/flox^*, *App ^flox/flox^Casr ^flox/flox^*, or *App^flox/flox^Gabbr1 ^flox/flox^Casr _flox/flox_* without PTH-Cre transgene. PTH-Cre, *Vdr ^flox/flox^*, and *App ^flox/flox^* mice were bred to generate ^PTC^*Vdr*^-/-^ and ^PTC^*Vdr*^-/-^*App*^-/-^ DKO mice. ^PTC^*Vdr*^-/-^ and *Mapt*^-/-^ mice were bred to generate ^PTC^*Vdr*^-/-^ //*Mapt*^-/-^ DKO mice, and control littermates. All transgenic mice were bred into C57/BL6 background for at least six generations and each line of gene knockout mice were inbred for at least three generations before being used for experiments at specified ages along with control littermates. Mouse genotypes were determined by PCR analysis of genomic DNA extracted from tail snips using primer sets reported previously. Blood was drawn from mice by cardiac puncture under anesthesia. Serum samples were analyzed for total calcium by an automated ACE Alera Clinical Chemistry bioanalyzer (Alfa Wassermann, Inc, West Caldwell, NJ, USA) and intact PTH by mouse PTH 1-84 EIA kits (Quidel Ortho 60-2305, San Diego, CA, USA), following manufacturer instructions. Our study examined male and female animals, and similar findings are reported for both sexes. All experiments were performed in accordance with the ARRIVE (Animal Research Reporting of In Vivo Experiments) guideline and approved by the Institutional Animal Care and Use Committee of the San Francisco Department of Veteran Affairs Medical Center.

### Ex vivo Study

#### PTG secretion assays

Live-cell interrogative assays for PTH secretion from human normal and adenoma specimens and the measurements of PTH secretion rate and Ca^2+^ set-point in mouse PTGs were performed as previously described (*7, 34*). Compounds tested on PTH secretion include Aβ_42_ (Aβ_1-42_) and its reverse peptide (Aβ_42-1_) (Bachem, Torrance, CA), custom sAPP_204-220_ (DDSDVWWGGADTDYADG) and its reverse peptide sAPP_220-204_, and CGP54626 (Tocris Bioscience, Minneapolis, MN). In some studies, after secretion assays, PTGs were compressed into 120 μm-thick discs using a pair of standard glass slide and coverslip with a Secure-SealTM spacer (S24737, Invitrogen by Thermo Fisher Scientific) during fixation by 4% paraformaldehyde. PTGs were then washed, stained, and imaged. Glandular volumes were calculated individually and used for statistical comparisons.

#### Immunohistochemistry

Deparaffinized sections of human or mouse PTGs were subjected to heat-induced antigen retrieval in 10 mM sodium citrate buffer pH 6.0 with 0.05% Tween-20, at 100°C for 10 min. Immunohistochemical detection of APP and Aβ_42_ in PTG sections were performed with a rabbit polyclonal anti-APP (CT695, Invitrogen) (2 μg/ml) and a custom-made mouse recombinant monoclonal anti-Aβ_42_ (murinized Aducanumab) (4 μg/ml), respectively, and corresponding Alex Fluor 488-conjugated secondary antibodies. Sections were then incubated with Texas Red-conjugated wheat germ agglutinin (WGA) and DAPI to label plasma membrane and nuclei, respectively. The specificity of anti-APP and anti-Aβ_42_ was confirmed by the lack of immunoreactivity in adjacent tissue sections subjected to non-immune IgG controls.

#### Proximity ligation assay (PLA)

This technique permits the detection and quantification of physical protein–protein interactions. When two selective antibodies are in proximity (< 30 nm) due to protein complex formation, complementary DNA strands attached to the secondary antibodies are allowed to be ligated, massively replicated, and visualized with fluorescence-conjugated DNA probes. To detect GABA_B1_R/CaSR complexes *in situ*, PLA were performed on sections of human PTGs using Duolink in Situ Detection kits (Sigma-Aldrich Corp., St Louis, MO) according to manufacturer’s instruction. Briefly, deparaffinized sections were incubated with a mouse monoclonal antibody against the N-terminus of GABA_B1_R (Abcam ab55051; 10 μg/ml) and a custom-made rabbit polyclonal antibody against the N-terminus of CaSR (VA609, ADDDYGRPGIEKFREEAEERDI; 4 μg/ml) at 4°C overnight, followed by sense and anti-sense oligonucleotide-conjugated anti-rabbit and anti-mouse IgG antibodies, respectively, at 37°C for 1 hour. DNA ligation was performed with DNA ligase (25U/ml) at 37°C for 1 hour, followed by DNA amplification with polymerase I (12.5U/ml) and nucleotides in the presence of Texas Red-conjugated DNA probes for 100 min at 37°C. Sections were then stained with Alex Fluor 477-conjugated mouse monoclonal anti-Aβ_42_ (Aducanumab, 4 μg/ml, 4°C overnight), washed, mounted with DAPI-containing mounting medium, imaged with a Zeiss Axio Imager 2 Microscopes (Carl Zeiss, Germany), and quantified using automated TissueQuest Analysis software (TissueGnositics USA, Ltd, Tarzana, CA). Total PLA activities in the regions of interest (ROIs) were divided by DAPI-positive cell number in the ROI to obtain mean PLA activity/cell.

#### NanoString nCounter assays

Total RNA was isolated from human parathyroid adenomas and PTGs of normal donors by an RNA STAT60 kit (Amsbio LLC, Cambridge, MA). A custom-made panel of 4-color, 6-spot optical barcoded DNA probes was used to assess the expression of 80 genes involved in parathyroid function, GPCR activation, and APP processing. 300 ng of total RNA was hybridized with barcoded reporter probes and biotinylated capture probes at 65°C for 16 hours. Hybridized transcript/probe complexes were immobilized onto streptavidin coated cartridges following manufacturer’s instruction. Abundance of transcripts was digitally counted by an nCounter Max analysis system (Bruker Spatial Biology, Seattle, WA). Transcript counts of genes of interest were normalized to housekeeping genes including Dnajc14, Gapdh, Rpl19, and Sdha using nSolver 4.0 software (Bruker Spatial Biology).

#### GeoMx Digital Spatial Profiling (DSP) protein assays

Five μm-thick FFPE sections were prepared from parathyroid adenomas and normal PTGs (table S1), deparaffinized, and subjected to heat-inducible antigen retrieval procedures (15 min in pressure cooker with 10 mM sodium citrate buffer pH 6.0, 0.05% Tween-20), then were incubated with fluorescent labeled antibodies [Cy5-conjugated mouse monoclonal anti-N-CaSR (3H8E9, ADDDYGRPGIEKFREEAEERDI; 5 ug/ml) and Texas Red-conjugated mouse monoclonal anti-TOM20 (F-10, Santa Cruz Biotechnology sc-17764; 2ug/ml)], together with photocleavable oligonucleotide-tagged profiling antibodies and nucleus stain Syto13 (500 nM) at 4°C for 16 hours. Next, slides were scanned at 20X magnification on the GeoMx Digital Spatial Profiler (Bruker Spatial Biology), and 3 to 5 regions of interest (ROI) were selected per tissue section. ROIs were chosen based on morphology markers (CaSR+/Tomm20-low/Syto13+ and CaSR+/Tomm20-high/Syto13+ for chief cell-enriched and oxyphil-enriched compartment, respectively). Subsequently, oligonucleotide tags were released from these compartments after exposure to 385 nm UV light and collected by microcapillaries in a 96-well plate. Released indexing oligonucleotides were dried down, resuspended in diethyl pyrocarbonate (DEPC)-treated water, hybridized to barcoded probes, and digitally counted using the nCounter MAX system (Bruker Spatial Biology). GeoMx software was used to normalize the digital counts using internal spike-in controls (ERCCs) and a panel of housekeeping proteins and to map counts back to tissue location yielding a spatially resolved digital profile of protein abundance. The 49 profiling antibodies were from Neural Cell Profiling Core module (GMX-PROCO-NCT-HNCP-12), Alzheimer’s Disease Pathology Module (GMX-PROMOD-NCTHADP-12), Alzheimer’s Disease Pathology Extended Module (GMX-PROMOD-NCTHADEP-12), and a custom GPCR module of antibodies to detect CaSR (MAb 1C12D7), GABA_B1_R (Abcam, ab264069, RabMAb EPR22954-47), and GABBR2 (Abcam, ab230136,RabMAb EP2411). Data analyses revealed indistinguishable protein profiles between oxyphils and chief cells in each tissue section, so they are combined in the reported data.

### In vitro Study

#### Live cell cAMP signaling

The rat parathyroid PTH-C1 cells, a generous gift from Dr. Maria Luisa Brandi, were cultured in DMEM/F12 (Corning Life Science, Glendale, AZ). PTH-C1 cells were seeded on 25 mm glass coverslips coated with collagen and transiently transfected with plasmids carrying cDNAs encoding CaSR, GABA_B1_R, and the intramolecular FRET cAMP sensor (epac1-CFP/YFP) using lipofectamine 3000 (Thermofischer Scientific) for 48–72 hours before experiments. Cells were immersed in HEPES buffer (137 mM NaCl, 5 mM KCl, 0.25 mM CaCl_2_, 1 mM MgCl_2_, 20 mM HEPES, 0.1% (w/v) bovine serum albumin (BSA), pH 7.4) in Attofluor cell chambers (Thermofischer Scientific) and mounted on a Nikon Ti-E inverted microscope (Nikon Corporation) with Perfect Focus System and equipped with an oil immersion 40× N.A. 1.30 Plan Apo objective and a moving stage (Nikon Corporation). Cells were pretreated with cholera toxin (CTx) for 1 hour to elevate basal cAMP, then changes in cAMP production were recorded by FRET changes following the sequential addition of CaCl_2_, Aβ_42_, and forskolin. Fluorescence emissions were recorded using a 480 ± 20 nm (for CFP) and 535 ± 15 nm (for YFP) filter set and collected simultaneously with a LUCAS EMCCD camera (Andor Technology) using a DualView 2 (Photometrics) with a beam splitter dichroic long pass (DCLP) of 505 nm. Fluorescence data were recorded from single cells using Nikon Element Software (Nikon Corporation). The FRET ratio for fluorescence emissions of CFP and YFP (F_YFP_/F_CFP_) for single cells was calculated and corrected for background, bleed-through, and photobleaching as previously described (*35*).

### In vivo Study

#### Generation of murinized aducanumab

A chimeric aducanumab monoclonal antibody was created by fusing the BIIB037 heavy and light chain variable regions obtained from the original patent (WO 20156/087944) with mouse IgG1 kappa heavy and light chain constant regions (*36*). Amino acid residues 1-124 of the heavy chain and amino acid residues 1-107 of the light chain were murine codon-optimized and joined to the constant regions of mouse IgG1 kappa heavy chain and light chain constant regions, respectively. Amino acid sequences can be found in the IMGT 2D structure database (*37*), under chain ID INN 9838 (https://imgt.org/3Dstructure-DB/cgi/details.cgi?pdbcode=9838&Part=Chain). cDNAs encoding the chimeric heavy and light chain peptides were cloned into the mammalian expression vector pCDNA3.4 and then co-transfected into CHO cells for antibody production. Secreted antibody was purified from cell culture supernatant using Protein G Sepharose chromatography. Purity was verified by mass spectrometry and SDS-PAGE. *In vivo* dosing was based on published studies (*38, 39*).

#### Calcimimetic and aducanumab regimen

24 MOA C57/B6 mice were intraperitoneally injected daily with cinacalcet (once daily of 5 mg/kg) or weekly with murinized aducanumab at low (once weekly of 10 mg/kg) or high (twice weekly of 20 mg/kg) dose, or in combination for 5 weeks. Sera were collected two hours after the last cinacalcet dose and/or three days after the last aducanumab injection, and sCa and sPTH were measured as described.

### Statistics

Comparisons between groups were subjected to statistical analysis using 2-tails Student’s t test for two groups or one-way or two-way ANOVA followed by Sidak’s for multiple comparisons using Prism 10 (GraphPad Software, Inc., La Jolla, CA, USA). Animal, organ culture, and cultured cell sample sizes were determined by a power analysis using the following parameters: standard deviation=5–10% depending on the assay, two-sided test, p value=0.05, power of the test=0.8. Multiple cohorts of mice were bred separately and studied to ensure reproducibility. Data from two groups were represented as mean ± standard error of the mean (s.e.m.). Significance was assigned for p value < 0.05. Ca^2+^-dose responses in suppression of PTH secretion were fitted with 5-knot Spline functions to extrapolate PTH_Max_ and Ca^2+^-setpoint using Prism 10.

## Data Availability

All data produced in the present study are available upon reasonable request to the authors.

## List of Supplementary Materials

Fig. S1 to S8

Table S1

Data files S1 (Excel files)

## Acknowledgments

We thank B. Bettler (University of Basel) and M.L. Brandi (University of Florence) for providing the floxed-Gabbr1 mice and PTH-C1 cells, respectively.

## Author contributions

Conceptualization: CLT, WC, JK, JAS, and DS

Methodology: CLT, WC, JK, JPV

Investigation: CLT, WC, ZC, NS, JK, TG, SS, JPV, and JAS

Visualization: CLT, WC, TG, JK, SS, JPV

Funding acquisition: WC, JK, JPV, JAS

Supervision: WC, JK, JPV, JAS

Writing – original draft: CLT, WC, JK, JPV

Writing – review & editing: CLT, WC, JK, TG, JPV, DS, and JAS

## Data and materials availability

All data are available in the main text or the supplementary materials.

## Supplementary Materials

**fig. S1.**
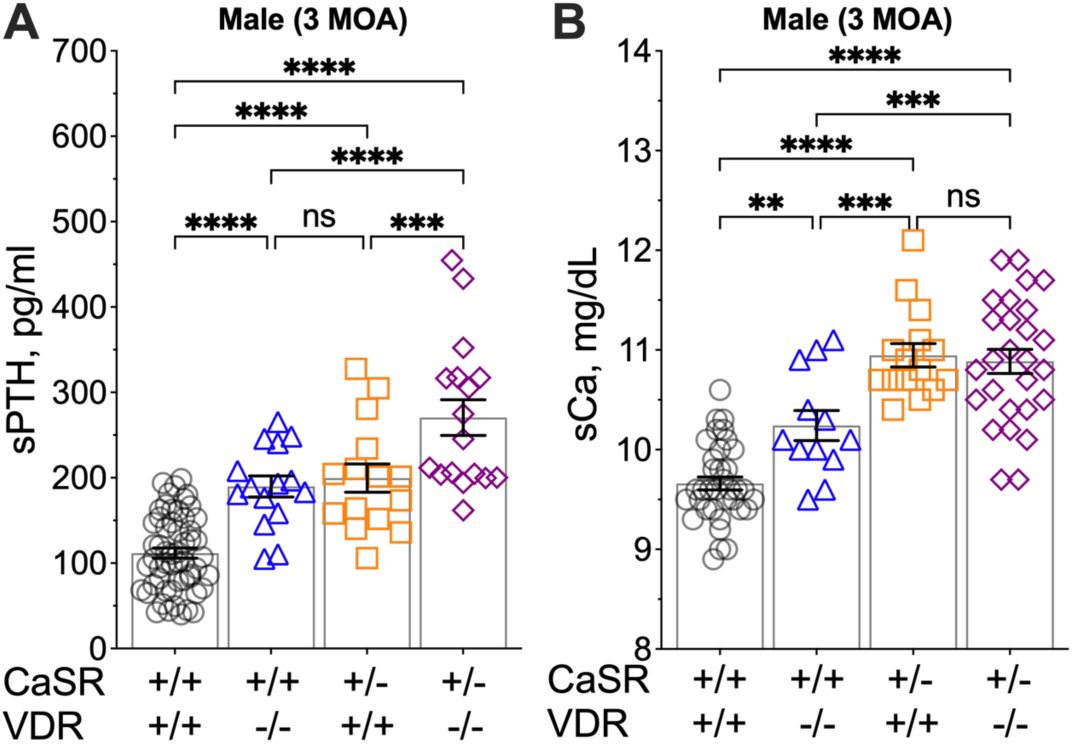
Hormonal and mineral status in mouse models of HPT. (**A**) Serum PTH (sPTH) and (**B**) total serum Ca^2+^ (sCa) of male ^PTC^*Vdr^-/-^* (blue triangles), ^PTC^*Casr^+/-^* (orange squares), ^PTC^*Vdr^-/-^ Casr^+/-^* (purple diamonds) mice and control littermates (Cont, black circles) at 3 MOA. Mean ± s.e.m. *n* = 10-32 mice. \*\**p* < 0.01, \*\*\**p* < 0.005, \*\*\*\**p* < 0.0001, and no significance (ns, *p* > 0.05) was determined by one-way ANOVA with Fisher’s LSD test using Prism 10 statistics software.

**fig. S2.**
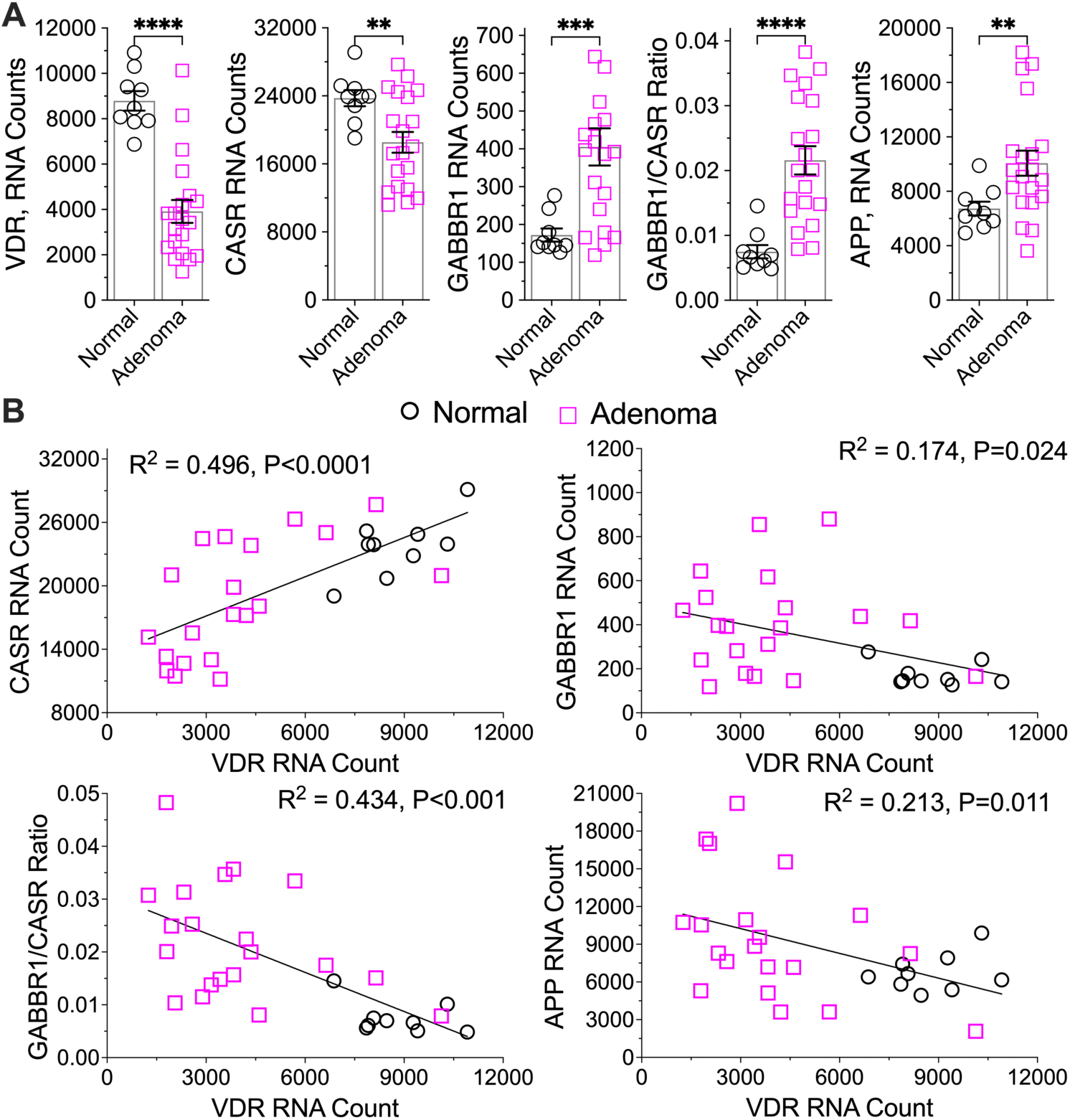
Decreased VDR RNA expression correlates with increased propensity of GABBR1/CaSR heterodimerization and APP expression in parathyroid adenomas from PHPT patients. (**A**) RNA abundance of VDR, CaSR, GABBR1 and APP in human parathyroid adenomas (pink squares, *n* = 17) and PTGs of normal donors (black circles, *n* = 7) was determined by NanoString nCounter gene expression assays. mean ± s.e.m. \*\**p* < 0.01, \*\*\**p* < 0.005, \*\*\*\**p* < 0.0001 by two-tailed student t-tests. (**B**) Correlation of each RNA level with VDR RNA level in parathyroid adenoma (pink squares) and normal PTGs (black circles) was performed by a linear regression model with derived Pearson coefficients (R^2^) and two-tailed *p* values at 95% confidence using Prism 10 statistics software.

**fig. S3.**
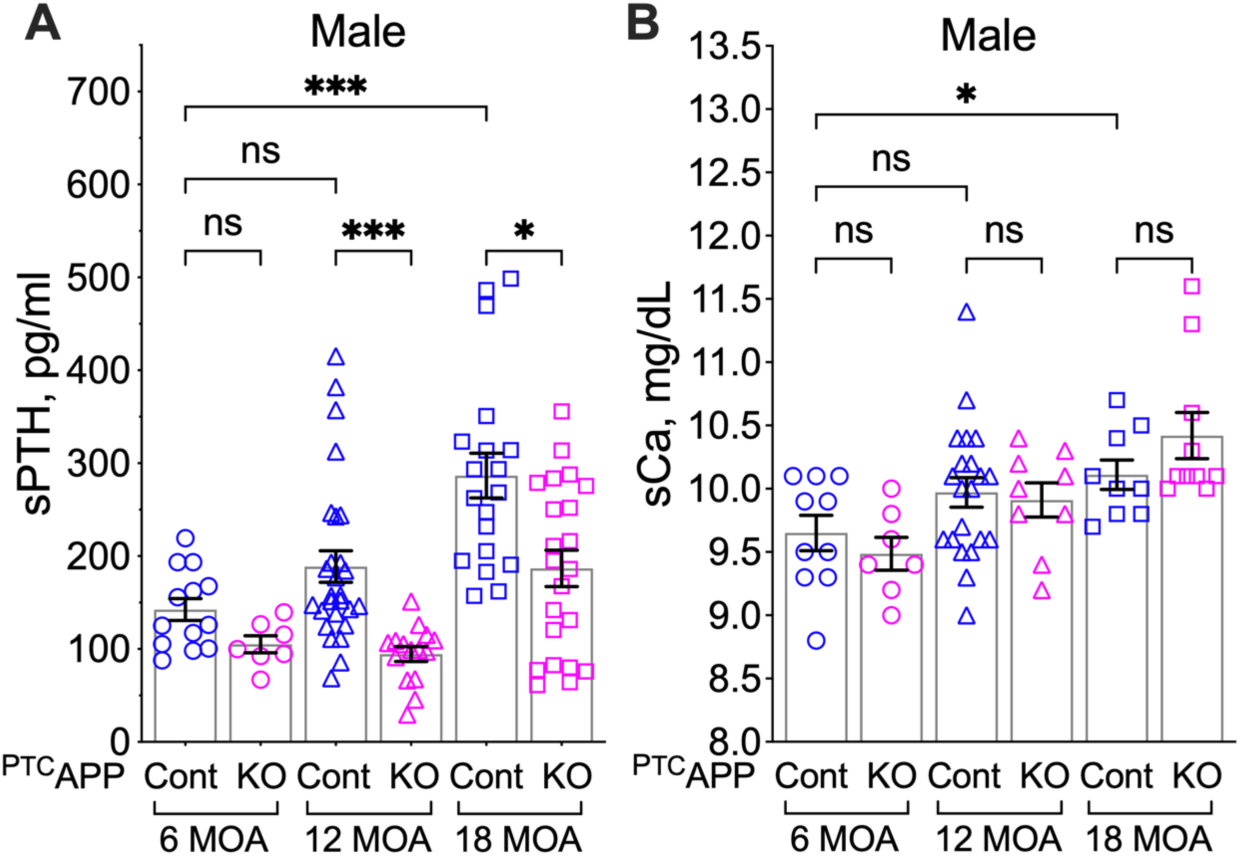
Impacts of PTC-specific *App* KO on hormonal and mineral status. (**A**) sPTH and (**B**) sCa levels of male *^PTC^App^-/-^* (KO, pink symbols) and *App*^fl/fl^ control (Cont, blue symbols) mice at 6 (circle), 12 (triangle), and 18 (square) MOA. Mean ± s.e.m. *n* = 7-26. ns (*p* > 0.05), \**p* < 0.05, \*\*\**p* < 0.005 vs control (two-way ANOVA with Sidak’s multiple comparisons test).

**fig. S4.**
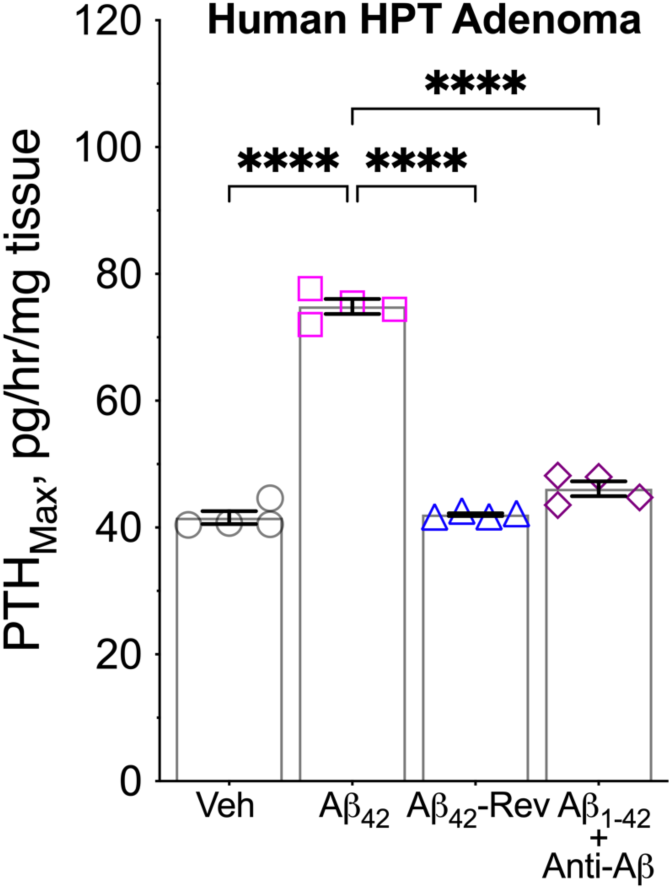
Effects of Aβ_42_ on PTH secretion in human parathyroid adenomas from HPT patients. Resected and sectioned parathyroid adenomas were incubated with vehicle (0.1% DMSO), Aβ_42-1_ (Aβ_42_-Rev, 200 nM), and Aβ_1-42_ (200 nM) with or without co-incubation with Aβ-neutralizing monoclonal antibody, Aducanumab (Anti-Aβ_42_, 20 μg/mL). Changes in maximal PTH secretion rate (PTH_Max_) on a per mg of tissue and per hour basis were determined from Ca^2+^-response curves. mean ± s.e.m., *n* = 4 gland sections for each treatment. \*\*\*\**p* < 0.001 (one-way ANOVA with Fisher’s LSD test).

**fig. S5.**
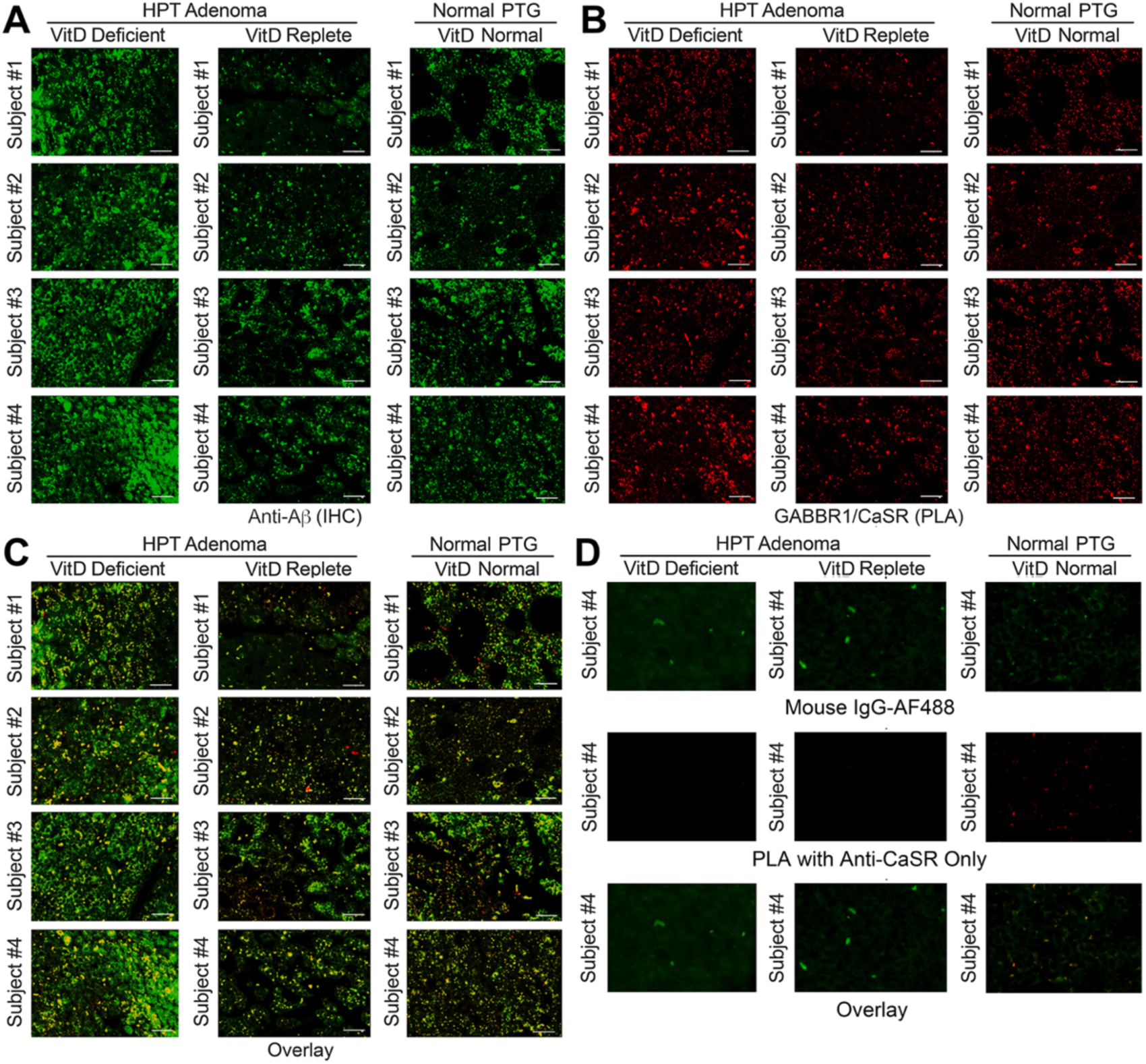
Vitamin D deficiency increases the expression and colocalization of Aβ_42_ with GABBR1/CaSR heterodimers. PTGs from 4 human subjects in each group of vitamin D deficient or repleted PHPT patients and normal donors were sequentially subjected to the proximity ligation assay (PLA) and immunohistochemistry (IHC) to detect endogenous GABBR1/CaSR heterodimers and Aβ_42_, respectively. (**A**) Aβ_42_ immunoreactivity was visualized with Alex Fluor 488 (green) and (**B**) GABBABR1/CaSR heterodimer with Texas Red (red) signals. (**C**) Colocalization of Aβ_42_ and GABBR1/CaSR were visualized by yellow signals in overlayed images. Scale bar: 125 µm. (**D**) The absence of fluorescent signals of receptor heteromerization or Aβ_42_ in PTGs treated with anti-CaSR alone in PLA or control IgG in IHC respectively.

**fig. S6.**
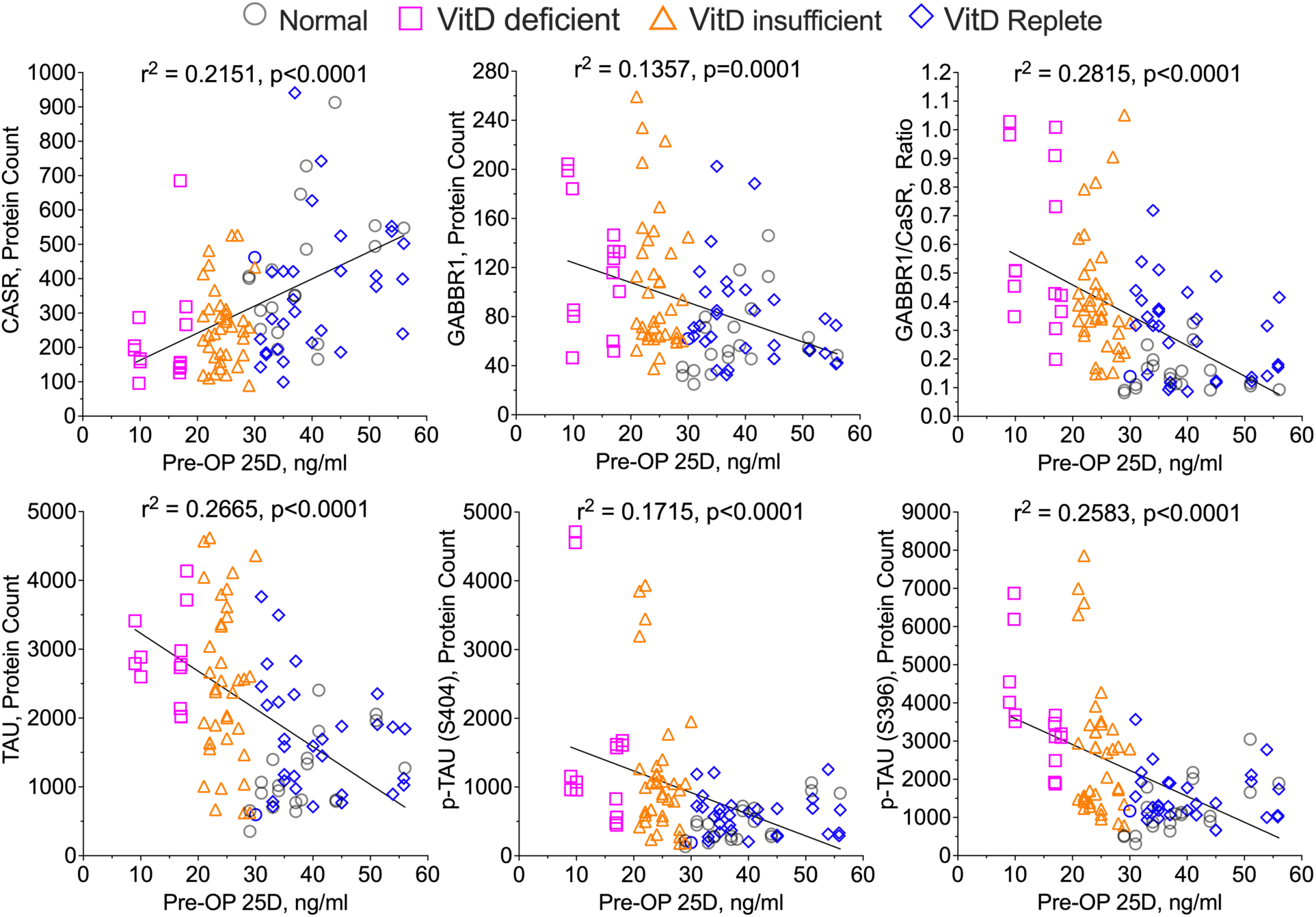
Pre-operative vitamin D levels inversely correlate with the propensity of GABBR1/CaSR heterodimerization and the expression and phosphorylation of Tau in human parathyroid. Protein expression in normal PTGs (grey circles) and parathyroid adenomas from PHPT patients with deficient (≤20 ng/ml, pink squares), insufficient (between 20-30 ng/ml, orange triangles), or replete (≥30 ng/ml, blue diamonds) pre-operative 25OH vitamin D levels were quantified by the NanoString GeoMx platform and correlated with pre-operative 25OH vitamin D levels using a linear regression model to derive the Pearson coefficients (R^2^) and two-tailed *p* values at 95% confidence using Prism 10 statistics software.

**fig. S7.**
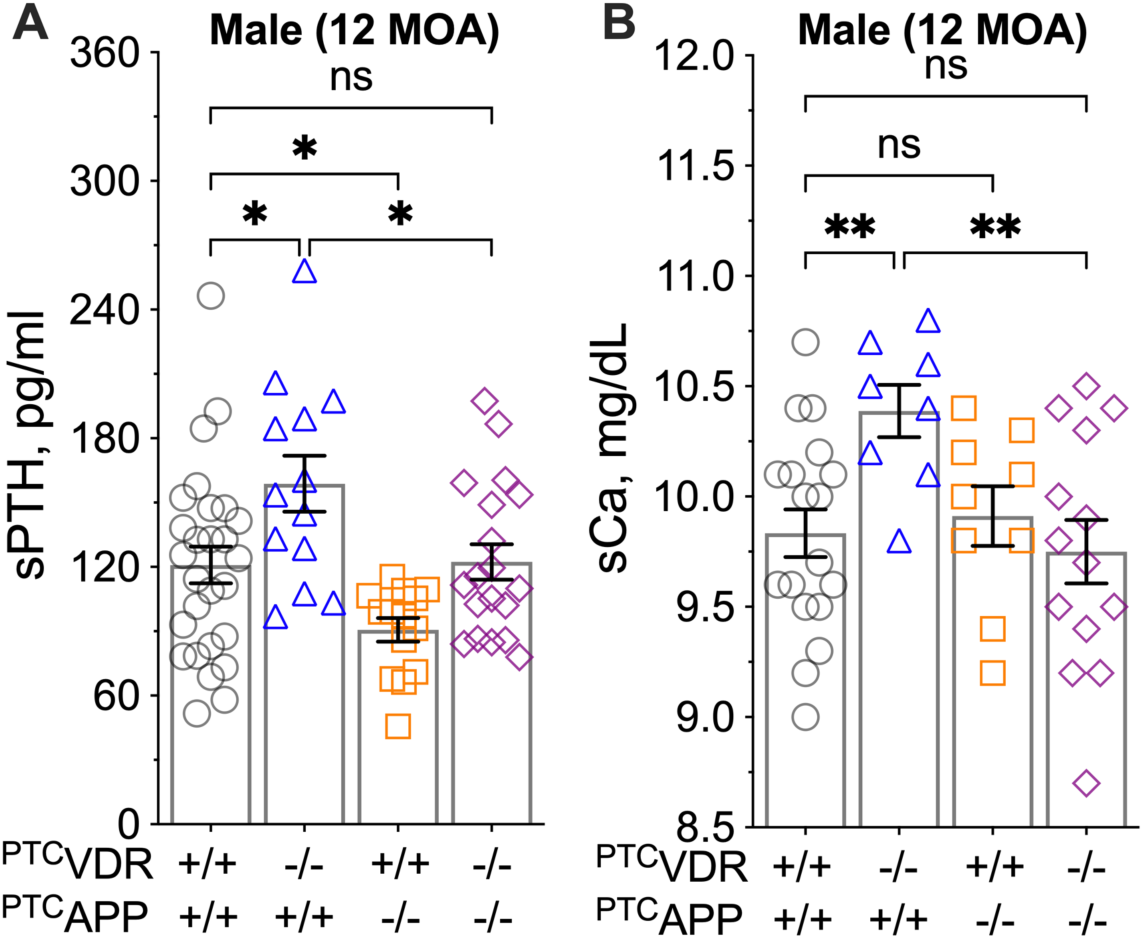
Impacts of PTC-specific *App* KO on hormonal and mineral status in mouse model of vitamin D deficiency. (**A**) sPTH and (**B**) sCa of male ^PTC^*Vdr*^-/-^ (blue triangles), ^PTC^*App*^-/-^ (orange squares), ^PTC^*Vdr*^-/-^*App*^-/-^ (purple diamonds) mice, and control littermates (*Vdr*^+/+^*App*^+/+^, grey circles) at 12 MOA. mean ± s.e.m., n = 8-28 mice for each genotype. ns (*p* > 0.05), \**p* < 0.05, \*\**p* < 0.01 (one-way ANOVA with Fisher’s LSD test).

**fig. S8.**
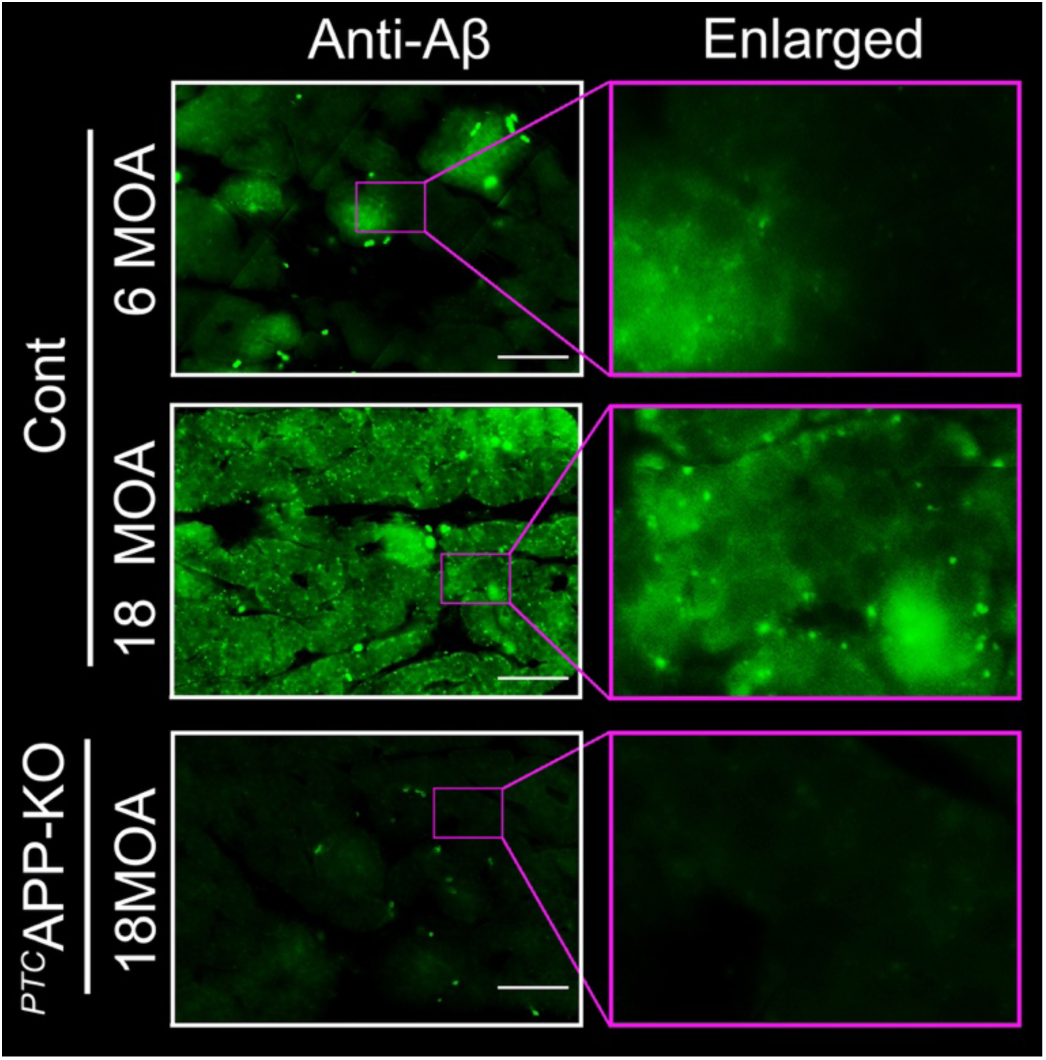
Increased expression of Aβ_42_ in PTGs of ageing mice. Immunohistochemical detections of endogenous Aβ_42_ in PTGs from 18 MOA *^PTC^App^-/-^* mice and *App*^fl/fl^ control littermates at 6 and 18 MOA. Aβ_42_ immunoreactivity was visualized with Alex Fluor 488 (green). Scale bar: 125 µm.

**table S1.** Demographics and serum chemistry of HPT patients and normal PTG donors. Vitamin D deficiency, insufficiency, and replete status are defined as pre-operative 25OHD levels ≤ 20 ng/ml, between 20-30 ng/ml, and ≥ 30 ng/ml, respectively, based on Institute of Medicine Guidelines (*34*). Mean ± s.e.m. *p*-value versus normal donors were determined by two-tailed Student’s t-test are indicated.

| Normal Donors |  |  |  |  |  | HPT: VitD Deficient |  |  |  |  |  |
| --- | --- | --- | --- | --- | --- | --- | --- | --- | --- | --- | --- |
| Sample ID | Sex | Age (years) | Pre-op 25D (ng/ml) | Pre-op PTH (pg/ml) | Pre-op sCa (mg/dL) | Sample ID | Sex | Age (years) | Pre-op 25D (ng/ml) | Pre-op PTH (pg/ml) | Pre-op sCa (mg/dL) |
| AHGP057RL | Female | 29-74 | 41 | ND | 10.4 | PTAS0020 | Female | 40-79 | 17 | 100 | 10.7 |
| AHBE267 |  |  | 39 |  | 9.1 | PTAS0064 |  |  | 18 | 45 | 10.8 |
| AHGY385LL |  |  | 51 |  | 8.2 | PTAS0104 |  |  | 9 | 119 | 10.9 |
| AHGN409RU |  |  | 44 |  | 9.6 | PTAS0126 |  |  | 10 | 260 | 10.7 |
| AGKC257 |  |  | 37 |  | 8.8 | PTAS0141 |  |  | 17 | 220 | 11.3 |
| AHHA246LU | Male |  | 55 |  | 10.2 | PTAS0094 |  |  | 17 | 62 | 10 |
| AHHA004LU |  |  | 38 |  | 10.1 | PTAS0128 |  |  | 18 | 88 | 10.2 |
|  |  |  |  |  |  | PTAS0057 |  |  | 9.8 | 300 | 10.9 |
| Mean |  | 44.6 | 43.6 |  | 9.5 | Mean |  | 59.1 | 14.5 | 149.3 | 10.7 |
| N |  | 7 | 7 |  | 7 | N |  | 8 | 8 | 8 | 8 |
| SD |  | 15.6 | 6.9 |  | 0.82 | SD |  | 13.57 | 4.07 | 96.78 | 0.41 |
|  |  |  |  |  |  | p value vs. Normal |  | 7.91E-02 | 3.14E-06 | ND | 7.01E-03 |
| HPT: VitD Insufficient |  |  |  |  |  | HPT: VitD Replete |  |  |  |  |  |
| Sample ID | Sex | Age (years) | Pre-op 25D (ng/ml) | Pre-op PTH (pg/ml) | Pre-op sCa (mg/dL) | Sample ID | Sex | Age (years) | Pre-op 25D (ng/ml) | Pre-op PTH (pg/ml) | Pre-op sCa (mg/dL) |
| PTAS0138 | Female | 37-80 | 21 | 112 | 10.7 | PTAS0084 | Female | 38-81 | 30 | 192 | 10.3 |
| PTAS0099 |  |  | 29 | 79 | 10.8 | PTAS0125 |  |  | 30 | 69 | 10.4 |
| PTAS0037 |  |  | 22 | 94 | 10.9 | PTAS0054 |  |  | 31 | 90 | 9.8 |
| PTAS0046 |  |  | 22 | 118 | 10.7 | PTAS0025 |  |  | 32 | 157 | 10.4 |
| PTAS0068 |  |  | 22 | 148 | 11 | PTAS0136 |  |  | 33 | 74 | 11 |
| PTAS0124 |  |  | 22 | 65 | 10.9 | PTAS0059 |  |  | 35 | 147 | 10.4 |
| PTAS0123 |  |  | 23 | 104 | 9.9 | PTAS0117 |  |  | 35 | 135 | 11.9 |
| PTAS0119 |  |  | 24 | 115 | 11.1 | PTAS0086 |  |  | 37 | 115 | 10.9 |
| PTAS0150 |  |  | 24 | 191 | 10.8 | PTAS0200 |  |  | 37 | 150 | 9.9 |
| PTAS0078 |  |  | 25 | 236 | 10.9 | PTAS0140 |  |  | 42 | 125 | 11.2 |
| PTAS0121 |  |  | 25 | 130 | 11.1 | PTAS0079 |  |  | 45 | 87 | 10 |
| PTAS0051 |  |  | 26 | 62 | 11 | PTAS0093 |  |  | 51 | 66 | 11.1 |
| PTAS0060 |  |  | 26 | 133 | 10.4 | PTAS0166 |  |  | 56 | 93 | 10.4 |
| PTAS0146 |  |  | 26 | 202 | 10.6 | PTAS0115 |  |  | 58 | 120 | 10.6 |
| PTAS0007 |  |  | 28 | 93 | 10.9 |  |  |  |  |  |  |
| PTAS0231 |  |  | 21 | 49 | 10.8 |  |  |  |  |  |  |
| PTAS0142 |  |  | 29 | 136 | 10.8 |  |  |  |  |  |  |
| PTAS0133 |  |  | 30 | 146 | 11.2 |  |  |  |  |  |  |
| Mean |  | 62.6 | 24.7 | 122.9 | 10.8 | Mean |  | 64.5 | 39.4 | 115.7 | 10.6 |
| N |  | 18 | 18 | 18 | 18 | N |  | 14 | 14 | 14 | 14 |
| SD |  | 10.68 | 2.89 | 49.65 | 0.30 | SD |  | 12.27 | 9.56 | 37.78 | 0.57 |
| p value vs. Normal |  | 2.17E-02 | 2.44E-04 | #DIV/0! | 4.75E-03 | p value vs. Normal |  | 3.71E-01 | 6.58E-08 | 3.74E-01 | 6.60E-01 |

